# Infection fatality rate of COVID-19 in community-dwelling populations with emphasis on the elderly: An overview

**DOI:** 10.1101/2021.07.08.21260210

**Authors:** Cathrine Axfors, John P A Ioannidis

## Abstract

**Objective:** This mixed design synthesis aimed to estimate the infection fatality rate (IFR) of Coronavirus Disease 2019 (COVID-19) in community-dwelling elderly populations and other age groups from seroprevalence studies. Protocol: https://osf.io/47cgb.

**Methods and analyses:** Eligible were seroprevalence studies done in 2020 and identified by any of four existing systematic reviews; with ≥1000 participants aged ≥70 years that presented seroprevalence in elderly people; that aimed to generate samples reflecting the general population; and whose location had available data on cumulative COVID-19 deaths in elderly (primary cutoff ≥70 years; ≥65 or ≥60 also eligible). We extracted the most fully adjusted (if unavailable, unadjusted) seroprevalence estimates. We also extracted age- and residence-stratified cumulative COVID-19 deaths (until 1 week after the seroprevalence sampling midpoint) from official reports, and population statistics, to calculate IFRs corrected for unmeasured antibody types. Sample size-weighted IFRs were estimated for countries with multiple estimates. Secondary analyses examined data on younger age strata from the same studies.

**Results:** Twenty-five seroprevalence surveys representing 14 countries were included. Across all countries, the median IFR in community-dwelling elderly and elderly overall was 2.9% (range 0.2%-6.9%) and 4.9% (range 0.2%-16.8%) without accounting for seroreversion (2.4% and 4.0%, respectively, accounting for 5% monthly seroreversion). Multiple sensitivity analyses yielded similar results. IFR was higher with larger proportions of people >85 years. Younger age strata had low IFR values (median 0.0013%, 0.0088%, 0.021%, 0.042%, 0.14%, and 0.65%, at 0-19, 20-29, 30-39, 40-49, 50-59, and 60-69 years even without accounting for seroreversion).

**Conclusions:** The IFR of COVID-19 in community-dwelling elderly people is lower than previously reported. Very low IFRs were confirmed in the youngest populations.

## INTRODUCTION

Most Coronavirus Disease 2019 (COVID-19) affect the elderly (1), and persons living in nursing homes are particularly vulnerable (2). Hundreds of seroprevalence studies have been conducted in various populations, locations, and settings. These data have been used and synthesized in several published efforts to obtain estimates of the infection fatality rate (IFR, proportion of deceased among those infected), and its heterogeneity (3-6). All analyses identify very strong risk-gradient based on age, although absolute risk values still have substantial uncertainty. Importantly, the vast majority of seroprevalence studies include very few elderly people (7). Extrapolating from seroprevalence in younger to older age groups is tenuous. Elderly people may genuinely have different seroprevalence. Ideally, elderly should be more protected from exposure/infection than younger people, although probably the ability to protect the elderly has varied substantially across countries (8). Moreover, besides age, comorbidities and lower functional status markedly affects COVID-19 death risk (9, 10). Particularly elderly nursing home residents accounted for 30-70% of COVID-19 deaths in high-income countries in the first wave (2), despite comprising <1% of the population. IFR in nursing home residents has been estimated to as high as 25% (11). Not separating residents of nursing homes from the community-dwelling may provide an average that is too low for the former and too high for the latter. Moreover, ascertainment and reporting of COVID-19 cases and deaths in nursing home populations show considerable variation across countries (2), with potentially heavy bearing on overall mortality, while community-dwelling elderly data may be less unreliable (especially in high-income countries). Finally, seroprevalence estimates reflect typically community-dwelling populations (enrollment of nursing home residents is scarce/absent in serosurveys).

Here we estimated the COVID-19 IFR in community-dwelling populations at all locations where seroprevalence studies with many elderly individuals have been conducted. Primary emphasis is on the IFR of the elderly. As a secondary analysis, we also explored the IFR of younger age-strata in these same studies.

## METHODS

### Information sources

We identified seroprevalence studies (peer-reviewed publications, official reports, or preprints) in four existing systematic reviews (3, 7, 12, 13) as for a previous project (14), using the most recent updates of these reviews and their respective databases as of November 23, 2021. All systematic reviews may miss some studies, despite their systematic efforts. In this project, the risk is minimized by using several existing systematic reviews of seroprevalence studies, each of them very meticulous. The protocol of this study was registered at the Open Science Framework (https://osf.io/47cgb) after piloting data availability in December 2020 but before extracting full data, communicating with local authorities and study authors for additional data and performing any calculations. Amendments to the protocol and their justification are described in Appendix Table 1.

**Table 1.**
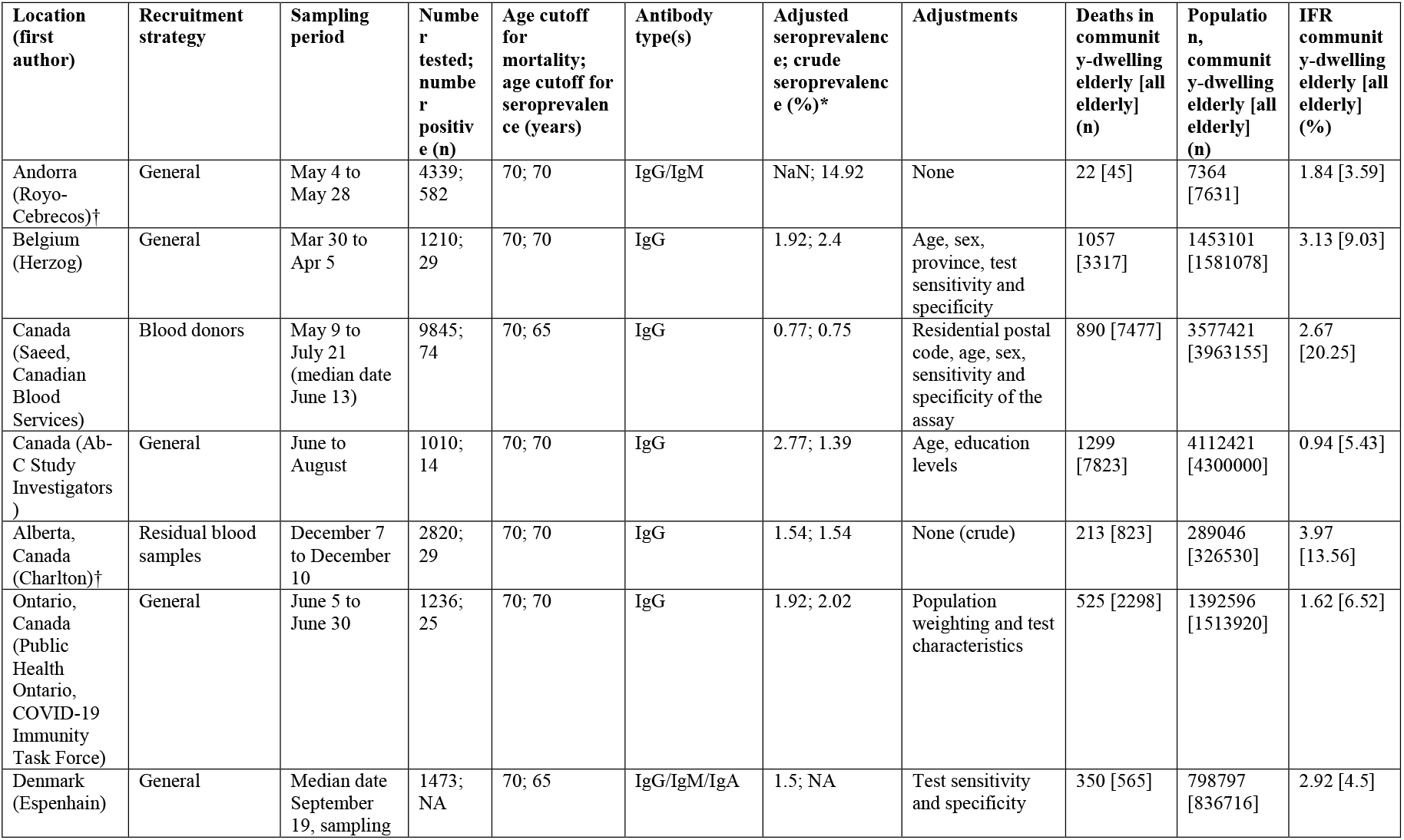
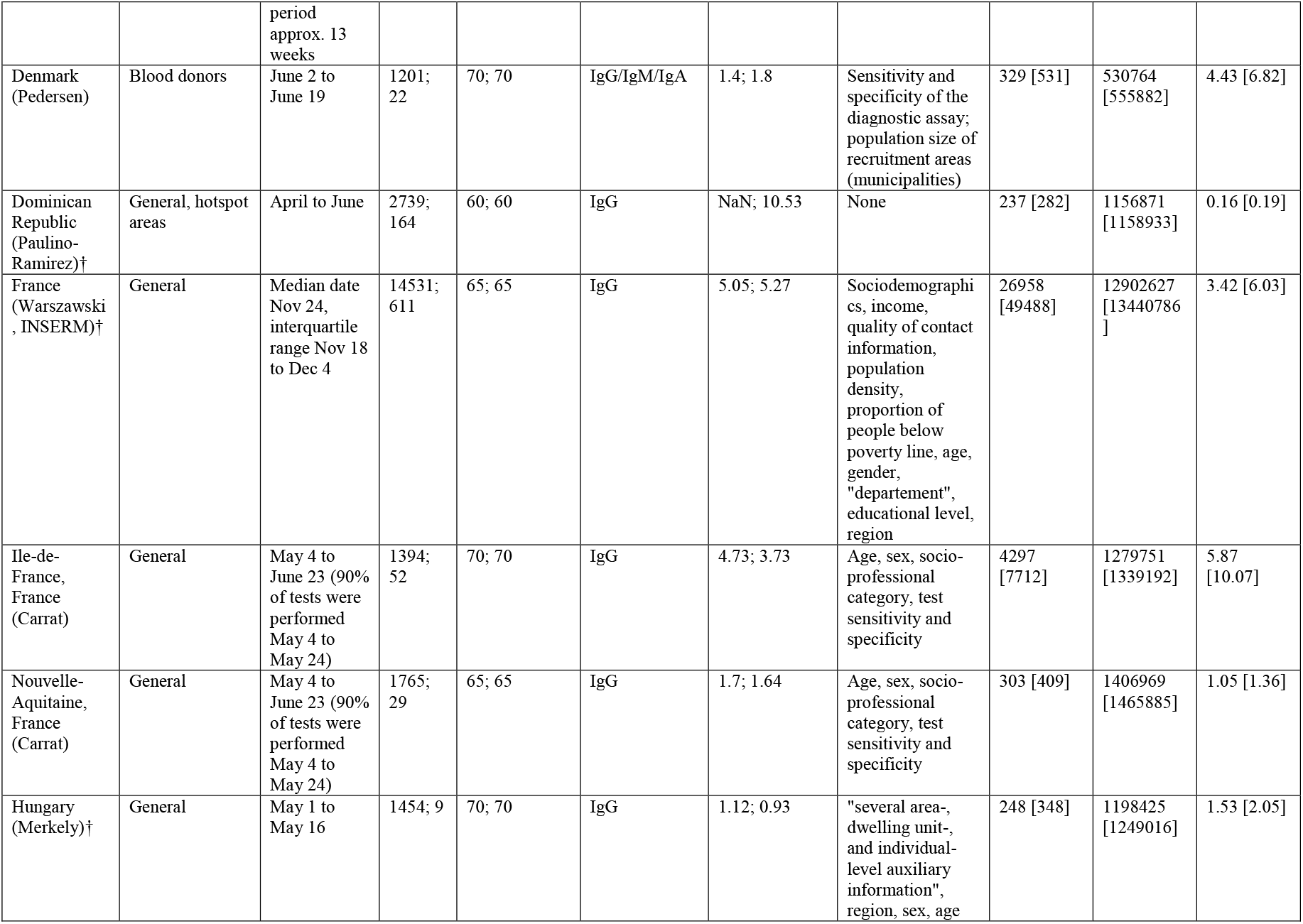
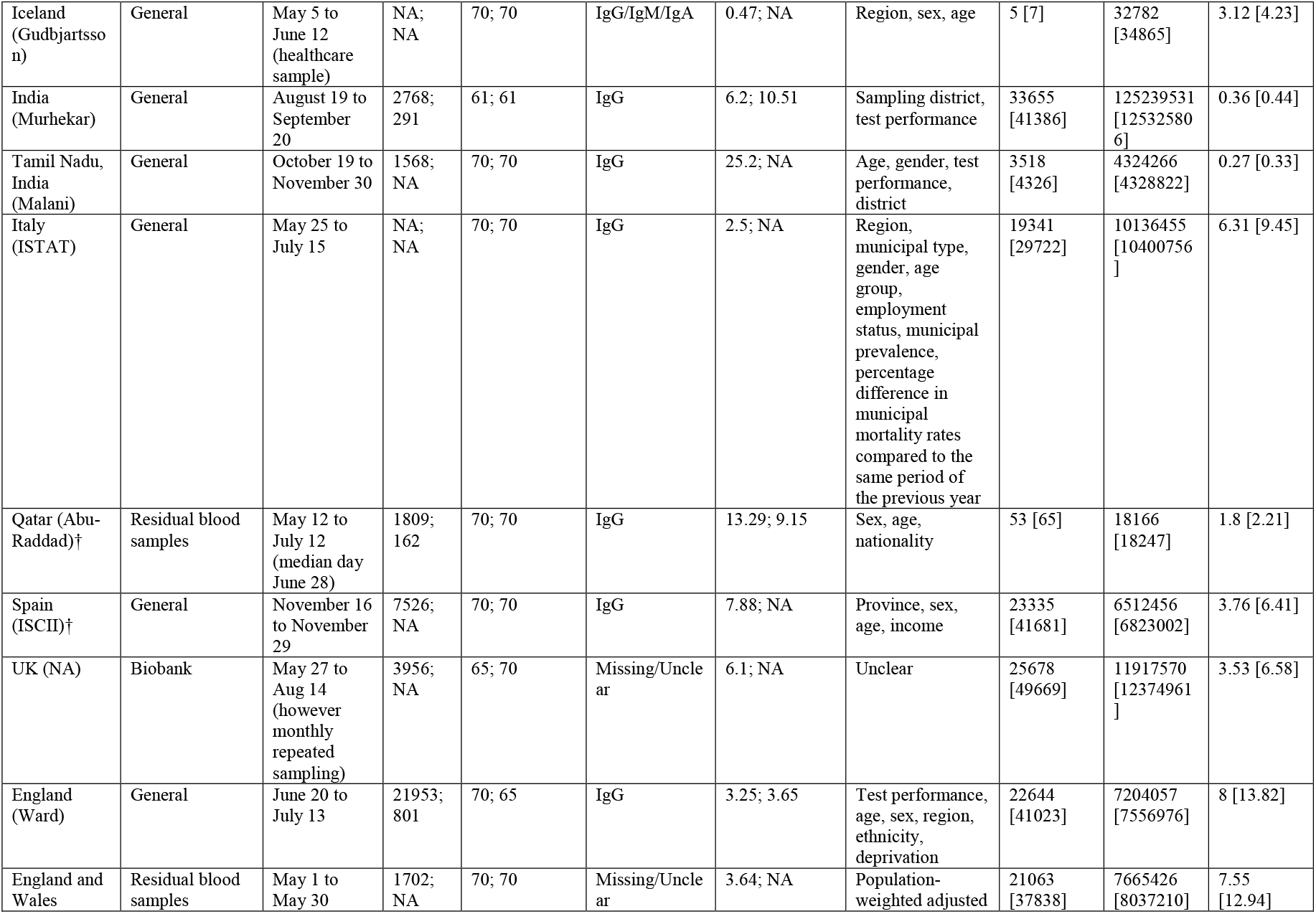
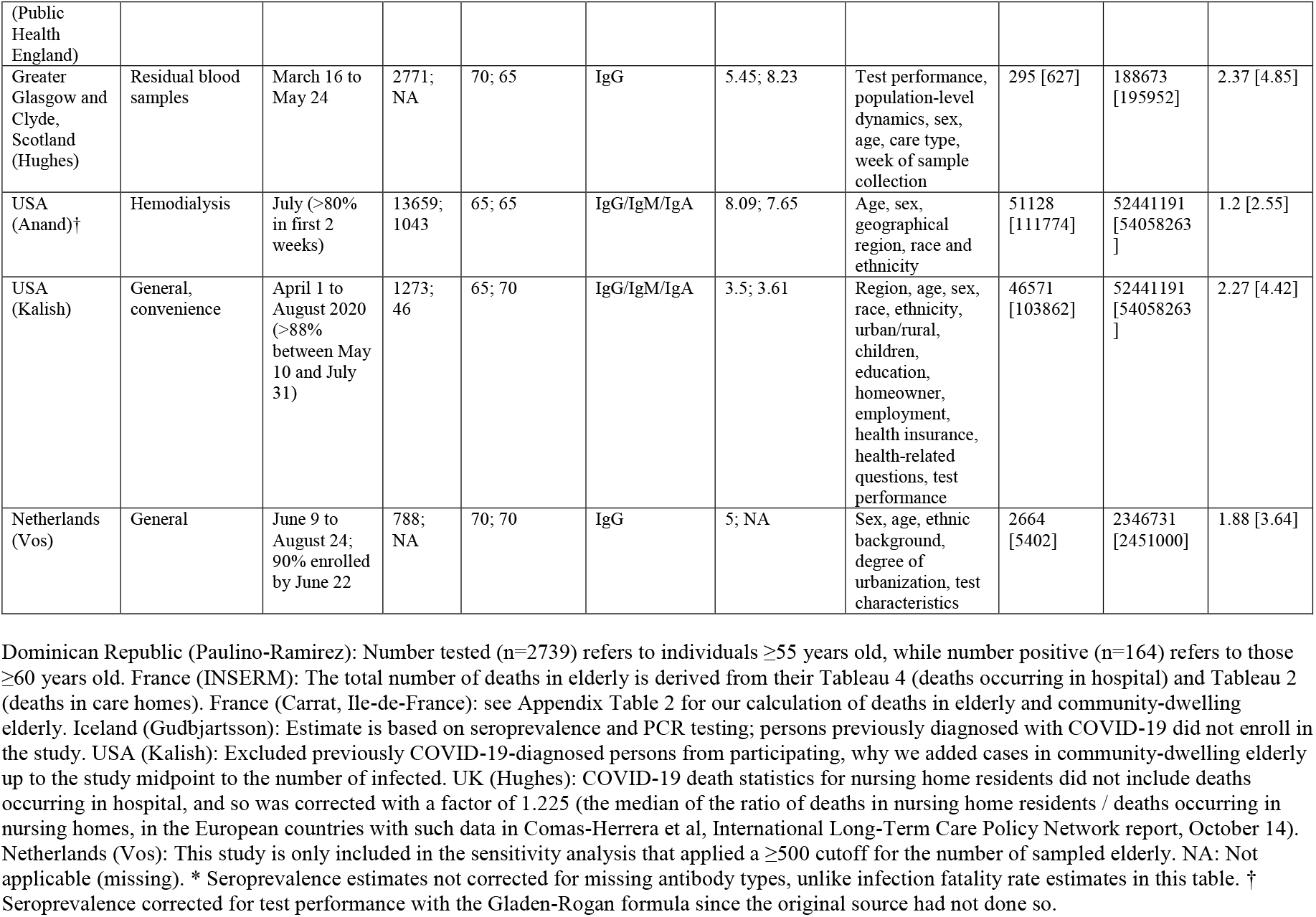
Included seroprevalence studies with estimates of seroprevalence in the elderly, COVID-19 deaths in the elderly and community-dwelling elderly, and corrected infection fatality rate

### Eligibility criteria

We included studies on SARS-CoV-2 seroprevalence that had sampled at least 1000 participants aged ≥70 years in the location and/or setting of interest, provided an estimate of seroprevalence for elderly people, explicitly aimed to generate samples reflecting the general population, and were conducted at a location for which there is official data available on the proportion of cumulative COVID-19 deaths among elderly (with a cutoff placed between 60-70 years; e.g., eligible cutoffs were ≥70, ≥65, or ≥60, but not ≥75 or ≥55). Besides general population samples we also accepted studies focusing on patient cohorts (including residual clinical samples), insurance applicants, blood donors, and workers (excluding health care workers and others deemed to have higher than average exposure risk, since these would tend to overestimate seroprevalence). USA studies were excluded if they did not adjust seroprevalence for race or ethnicity, since these socio-economically related factors associate strongly with both study participation (15, 16) (blood donation, specific jobs, and insurance seeking) and COVID-19 burden (17-19). Following comments from peer-reviewers, we have added another exclusion criterion: crude seroprevalence being less than 1-test specificity and/or the 95% confidence interval of the seroprevalence going to 0% (since the seroprevalence estimate would be extremely uncertain). We focused on studies sampling participants in 2020, since IFRs in 2021 may be further affected by wide implementation of vaccinations that may substantially decrease fatality risk and by other changes (new variants and better treatment). Following comments from peer-reviewers we also explored different eligibility criteria in two sensitivity analyses: (a) focusing only on explicitly general population samples based on the SeroTracker categories of “Household and community samples” and “Multiple general populations” and thus excluding for example patient cohorts, insurance applicants, blood donors, and workers, and (b) including only explicitly national-level general population studies without high risk of bias and with at least 500 participants aged ≥70 years. We applied the risk of bias assessments reported by the SeroTracker team (based on the Joanna Briggs Institute Critical Appraisal Tool for Prevalence Studies) (20). Two authors reviewed records for eligibility. Discrepancies were solved by discussion.

### Data extraction

CA extracted each data point and JPAI independently verified the extracted data. Discrepancies were solved through discussion. For each location, we identified the age distribution of cumulative COVID-19 deaths and chose as primary age cutoff the one closest to 70, while placed between 60-70 years (e.g., ≥70, ≥65, or ≥60).

Similar to a previous project (3), we extracted from eligible studies information on location, recruitment and sampling strategy, dates of sample collection, sample size (overall and elderly group), and types of antibody measured (immunoglobulin G (IgG), IgM and IgA). We also extracted, for the elderly stratum, the estimated unadjusted seroprevalence, the most fully adjusted seroprevalence, and the factors considered for adjustment. Antibody titers may decline over time. E.g. a modelling study estimated 3-4 months average time to seroreversion (21). A repeated measurements study (22) suggests even 50% seroreversion within a month for asymptomatic/oligosymptomatic patients, although this may be an over-estimate due to initially false-positive antibody results. To address seroreversion, if there were multiple different time points of seroprevalence assessment, we selected the one with the highest seroprevalence estimate. If seroprevalence data were unavailable as defined by the primary cutoff, but with another eligible cutoff (e.g., ≥70, ≥65, or ≥60), we extracted data for that cut-off.

Population size (overall, and elderly) and numbers of nursing home residents for the location were obtained from multiple sources (see Appendix Table 2).

Cumulative COVID-19 deaths overall and in the elderly stratum (using the primary age cutoff) for the relevant location were extracted from official reports. The total number, i.e., confirmed and probable, was preferred whenever available. We extracted the accumulated deaths until 1 week after the midpoint of the seroprevalence study period (or the closest date with available data) to account for different delays in developing antibodies versus dying from infection (23, 24). If the seroprevalence study claimed strong arguments to use another time point or approach, while reporting official statistics on the number of COVID-19 deaths overall and in the elderly population, we extracted that number instead.

The proportion of cumulative COVID-19 deaths that occurred among nursing home residents for the relevant location and date was extracted from official sources or the International Long Term Care Policy Network (ILTCPN) report closest in time (2, 25). We defined community-dwelling individuals by excluding persons living in institutions. Types of institutions used for elderly in various countries differed in nature and in the frailty of individuals residing there. For each location, we extracted available definitions of institutions. We preferred numbers recorded per residence status, i.e., including COVID-19 deaths among nursing home residents occurring in hospital. If the latter were unavailable, we calculated the total number of deaths in nursing home residents with a correction (by multiplying with the median of available ratios of deaths in nursing homes to deaths of nursing home residents in the ILTCPN 10/14/2020 report (2) for countries in the same continent). We considered 95%, 98%, and 99% of nursing home residents’ deaths to have occurred in people ≥70 years, ≥65 years and ≥60 years, respectively (26). For other imputations, see the online protocol.

### Missing data

We communicated with the authors of the seroprevalence study and with officers responsible for compiling the relevant official reports to obtain missing information or when information was available but not for the preferred age cut-offs. Email requests were sent, with two reminders to non-responders.

### Calculated data variables

#### Infected and deceased community-dwelling elderly

The number of infected people among the community dwelling elderly for the preferred date (1 week after the midpoint of the seroprevalence study period) was estimated by multiplying the adjusted estimate of seroprevalence and the population size in community-dwelling elderly. We preferred the most adjusted seroprevalence estimates. Following a suggestion from peer-reviewers, whenever no adjustment was made for test performance, we adjusted the estimates for test performance using the Gladen-Rogan formula (27). Moreover, we applied a non-prespecified correction for studies that excluded persons with diagnosed COVID-19 from sampling, primarily by using study authors’ corrections, secondarily by adding the number of identified COVID-19 cases in community-dwelling elderly for the location up to the seroprevalence study midpoint.

The total number of fatalities in community-dwelling elderly was obtained by total number of fatalities in elderly minus those accounted for by nursing home residents in the elderly stratum. If the elderly proportion or nursing home residents’ share of COVID-19 deaths were only available for another date than the preferred one, we assumed that the proportions were stable between the time points.

#### IFR estimation

We present IFR with corrections for unmeasured antibodies (as previously described (3)) as well as uncorrected. When only one or two types of antibodies (among IgG, IgM, IgA) were used in the seroprevalence study, seroprevalence was corrected upwards (and inferred IFR downwards) by 10% for each non-measured antibody (28). We added a non-prespecified calculation of 95% confidence intervals (CIs) of IFRs based on extracted or calculated 95% CIs from seroprevalence estimates (Appendix Table 1). No further factors were introduced in the calculation. CI estimates should be seen with caution since they depend on adequacy of seroprevalence adjustments, and do not consider other types of uncertainty (e.g., regarding mortality statistics).

### Synthesis of data

Statistical analyses were done using R version 4.0.2 (29). Similar to a previous overview of IFR-estimating studies (3), we estimated the sample size-weighted IFR of community-dwelling elderly for each country and then estimated the median and range of IFRs across countries. As expected, there was extreme heterogeneity among IFR estimates (I^2^ = 99.2%), thus weighted meta-analysis averages are not meaningful (30, 31).

We explored a seroreversion correction of the IFR by X^m^-fold, where m is the number of months from the peak of the first epidemic wave in the specific location and X is 0.99, 0.95, and 0.90 corresponding to 1%, 5%, and 10% relative monthly rate of seroreversion (21, 22, 32). We also added a non-prespecified sensitivity analysis to explore the percentage increase in the cumulative number of deaths and IFR, if the cutoff was put two weeks (rather than 1 week) after the study midpoint.

We expected IFR would be higher in locations with a higher share of people ≥85 years old among the analyzed elderly stratum. Estimates of log_10_IFR were plotted against the proportion of people ≥85 years old among the elderly (for population pyramid sources see Appendix Table 2).

#### Added secondary analyses

IFR in younger age-strata has become a very important question since we wrote the original protocol and the studies considered here offered a prime opportunity to assess IFR also in younger age strata. Among the included studies, whenever there were seroprevalence estimates and COVID-19 mortality data available for younger age groups, we complemented data extraction for all available age strata. Studies were excluded if no mortality data were available for any age stratum of maximum width 20 years and maximum age 70 years. We used the same time points as those selected for the elderly data. We included all age strata with a maximum width of 20 years and available COVID-19 mortality information. We corresponded the respective seroprevalence estimates for each age stratum with eligible mortality data. Consecutive strata of 1-5 years were merged to generate 10-year bins. For seroprevalence estimates we used the age strata that most fully covered the age bin for which mortality data were available; for the youngest age groups seroprevalence data from the closest available group with any sampled persons ≤20 years were accepted. E.g. for Ward et al (33), eligible age strata were 0-19 (paired with seroprevalence data for 20-24), 20-24, 25-34, 35-44, 45-54, 55-64. Population statistics for each analyzed age bin were obtained from the same sources as for the elderly. For age strata with multiple estimates from the same country, we calculated the sample size-weighted IFR per country before estimating median IFRs across locations for age groups 0-19, 20-29, 30-39, 40-49, 50-59, and 60-69 years. IFR estimates were placed in these age groups according to their midpoint, regardless of whether they perfectly matched the age group or not, e.g. an IFR estimate for age 18-29 years was placed in the 20-29 years group. As for the main analysis, following a suggestion from peer-reviewers, whenever no adjustment was made for test performance, we adjusted the estimates for test performance using the Gladen-Rogan formula (27).

### Ethics approval

Not applicable to this study.

### Patient and Public Involvement

There was no involvement of patients nor the public in this research.

## RESULTS

### Seroprevalence studies

By November 23, 2021, 3138 SARS-CoV-2 seroprevalence reports were available in the four systematic reviews. Screening and exclusions are shown in Appendix Figure 1 and Appendix Table 3. Twenty-four seroprevalence studies were included, one of which contained two separate surveys.

**Figure 1.**
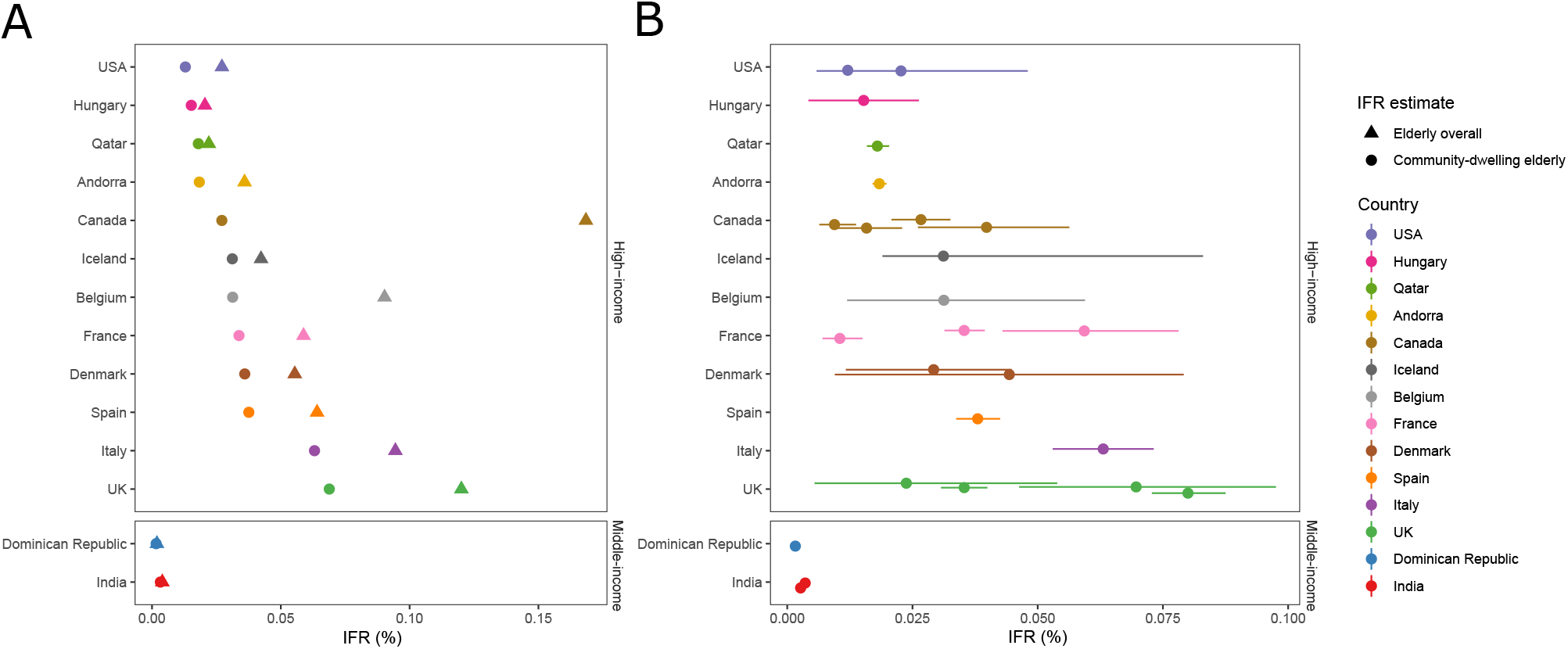
Infection fatality rates (IFRs) in elderly, corrected for unmeasured antibody types. **(A)** Countries’ IFRs in community-dwelling elderly and elderly overall. **(B)** IFRs in community-dwelling elderly with 95% confidence intervals based on individual seroprevalence estimates and their uncertainty. If multiple seroprevalence studies were available for the same country, we calculated the sample size-weighted IFR. As per above, the 95% CIs do not take into account other sources of uncertainty than those adjusted by the seroprevalence study authors (except adding an adjustment for test performance as per the Gladen-Rogan formula for those that had not already adjusted for test performance), and should be interpreted as conservative. Primarily, 95% confidence intervals are direct extractions from the seroprevalence studies. For studies that did not report 95% confidence intervals, we complemented with a calculation using the number of sampled and seropositive elderly individuals. For those that provided adjusted estimates for age brackets (e.g., 70-79, 80-89, and 90+), we combined estimates for each study using a fixed effects inverse variance meta-analysis (of arcsine transformed proportions) to obtain 95% CIs. Asymmetry to point estimates may be observed for these cases, since point estimates were calculated by multiplying age bracket seroprevalence by the corresponding population count (which is preferable, since it takes into account population distribution).

Table 1 shows for each eligible study the sampling period, sample size tested and positive, age cut-offs for the elderly group, antibody type(s), seroprevalence estimates, types of adjustments, number of deaths, and IFR estimates. The 25 seroprevalence surveys (Table 1) (33-57) represented 14 countries (Americas n=7, Asia n=3, Europe n=15). Only three studies were conducted in middle-income countries (one in Dominican Republic, two in India) and the other 22 in high-income countries. Seventeen studies targeted general population participants, 2 enrolled active or former blood donors, respectively (36, 40), 1 biobank participants (53), 1 hemodialysis patients (56), and 4 used residual blood samples (39, 50, 54, 55). Three studies excluded upfront persons with previously diagnosed COVID-19 from participating in their sample (46, 50, 57). Mid-sampling points ranged from April 2020 to December 2020. Sampling had a median length of 5.4 weeks (range 3 days to 5 months). The median number of elderly individuals tested was 1809 (range 1010-21953). Median seroprevalence was 3.5% (range 0.47%-25.2%). Adjusted seroprevalence estimates were available for 23/25 surveys.

### Mortality and population statistics

COVID-19 deaths and population data among elderly at each location are shown in Table 1 (for sources, see Appendix Table 2). The proportion of a location’s total COVID-19 deaths that happened among elderly had a median of 53% (range 51%-62%) in middle-income countries and 86% (range 51%-93%) in high-income countries. The proportion of a location’s total COVID-19 deaths that occurred in nursing home residents was imputed for middle-income countries, and had a median of 43% (range 20%-85%) in in high-income countries with available data (for Qatar, the number was imputed). One study (55) included only COVID-19 deaths that occurred in nursing homes and was corrected to reflect also the deaths among nursing home residents occurring in hospitals. Among the population, the elderly group comprised a median of 9% (range 6%-11%) in middle-income countries and 14% (range 0.6%-24%) in high-income countries. People residing in nursing homes were 0.08-0.20% of elderly in middle-income countries and a median of 4.8% (range 0.5%-12.8%) in high-income countries.

### Additional data contributed

Additional information was obtained from authors and agencies on four studies for seroprevalence data (35, 38, 45, 56); three studies for mortality data (34, 35, 38); two studies for population data (34, 35); and five excluded studies (clarifying non-eligibility).

### Calculated IFRs

For 6 countries with more than one IFR estimate available sample size-weighted average IFRs were calculated. In 14 countries, IFRs in community-dwelling elderly (Figure 1, Table 1) had a median of 2.9% (range 0.2%-6.9%). In two middle-income countries, IFR was 0.2% and 0.3%, versus a median of 3.1% (range 1.3%-6.8%) in 12 high-income countries. Figure 1 also shows 95% CIs for IFRs based on 95% CIs for seroprevalence estimates. For 9 studies, 95% CIs were direct extractions from the seroprevalence studies themselves, while complementary calculations were performed for the others as described in Appendix Table 1. For 8 studies, seroprevalence estimates were corrected for test performance using the Gladen-Rogan formula (Appendix Table 4). Median IFR in all elderly for all 14 individual countries was 4.9% (range 0.2%-16.8%). In the 2 middle-income countries, IFR in all elderly was 0.2% and 0.4% and in 12 high-income countries the median was 5.7% (range 2.1%-16.8%).

Sensitivity analyses exploring different rates of seroreversion appear in Appendix Table 5. For the scenario with 5% relative monthly seroreversion, median IFR in community-dwelling elderly was 2.4% (range 0.2%-5.9%) across all countries (0.1% and 0.3% in 2 middle-income countries, and 2.6% in 12 high-income countries); corresponding median IFR in all elderly was 4.0% (range 0.2%-14.8%). For the sensitivity analysis that explored the percentage increase in IFR if a later cutoff was used for cumulative deaths (two weeks after study midpoint), data were available for 22/25 seroprevalence surveys. There was a median relative increase of 4%, and median IFR in community-dwelling elderly became 3.0% (Appendix Table 6).

In additional analyses proposed by peer-reviewers, the median IFR was 2.9% when focusing only on explicitly general population samples (17 studies on 12 countries) and it was 3.1% among high-income countries (22 studies on 12 countries). The median IFR was 2.6% including explicitly national-level general population studies with at least 500 participants aged ≥70 years (18 studies on 13 countries) (screening and exclusions are shown in Appendix Figure 2). The median IFR was 2.8% upon excluding 4 studies where the selected time point with highest seroprevalence was not the latest available (seroprevalence had declined in the latest timepoint) (33, 36, 42, 54).

**Figure 2.**
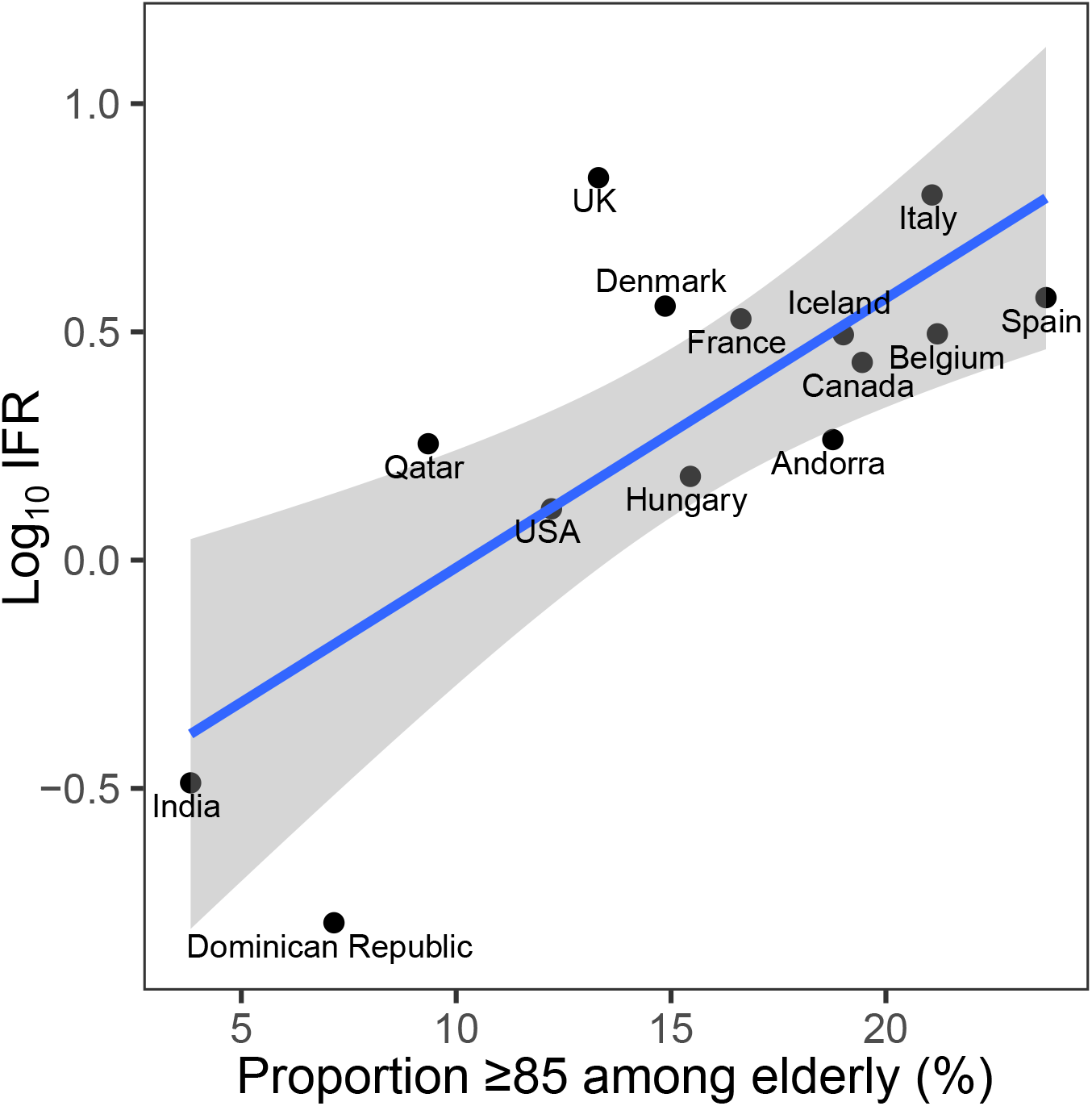
Infection fatality rate in community-dwelling elderly, corrected for unmeasured antibody types, plotted against the proportion of people ≥85 years old among the elderly. Log_10_ IFR: logarithm (with base 10) of the infection fatality rate. The “elderly” group is defined by the primary cutoff for each location. E.g. for Belgium 3% of the population is ≥85, and 13.6% of the population is ≥70, thus the proportion is 3/13.6. Imputation done for regional data: Denmark (3/5 regions), and Tamil Nadu, India, with country-level proportion of persons ≥85 years old among elderly.

### IFR in the elderly and proportion >85 years

There was steeply increasing IFR with larger proportions of people ≥85 years old (Figure 2). A regression of logIFR against the proportion of people ≥85 years old had a slope of 0.06 (p=0.002), and suggested an IFR in community-dwelling elderly of 0.49%, 0.98%, and 3.90% when the proportion of people >85 in the elderly group was 5%, 10%, and 20%, respectively.

### IFR in younger age-strata

We could extract data and calculate IFR on another 97 age-strata observations from 21/25 seroprevalence surveys (three had no mortality data for any eligible non-elderly age stratum (34, 40, 46) and one sampled no individuals <65 years of age (54)). The 21 surveys came from 12 countries. For the age group 0-19 years, only six studies had sampled participants for seroprevalence in the corresponding age group (33, 38, 39, 41, 49, 52); for the other studies, the closest available age group was used. Across all countries (Figure 3), the median IFR was 0.0013%, 0.0088%, 0.021%, 0.042%, 0.14%, and 0.65%, at 0-19, 20-29, 30-39, 40-49, 50-59, and 60-69 years, using data from all 12 countries for all age strata except 60-69, where data from 8 countries were used. Appendix Figure 3 visualizes these estimates against other, previously published evaluations of age-specific IFR.

**Figure 3.**
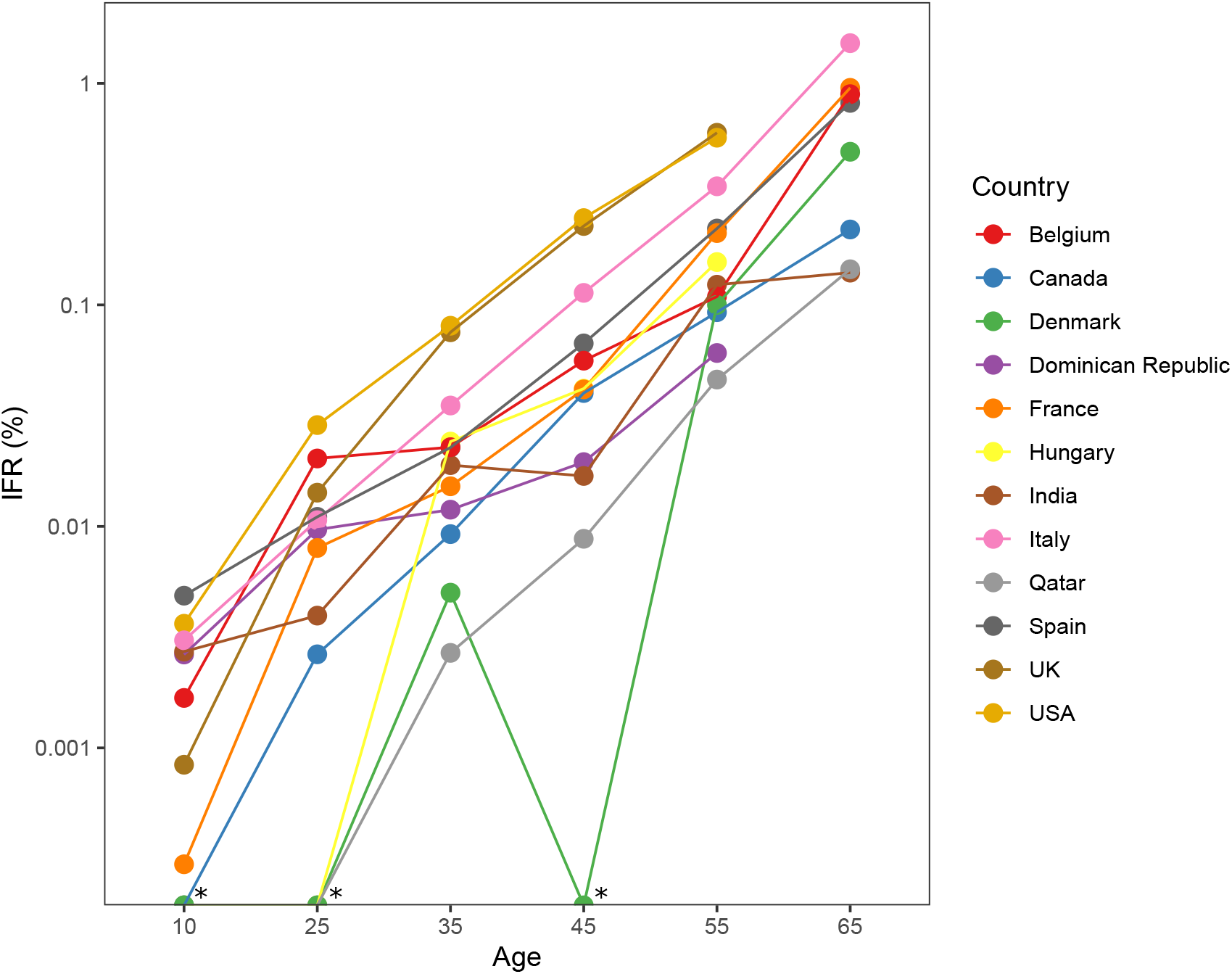
Infection fatality rates in younger age groups derived from included seroprevalence studies. IFRs are corrected for unmeasured antibody types. Sample size weighted IFRs were calculated for countries with multiple estimates available. * Infinite values produced by zero death counts.

## DISCUSSION

The IFR of COVID-19 in elderly was found to vary widely at locations where seroprevalence studies have enrolled many elderly individuals. IFR in community-dwelling elderly was consistently lower than in elderly overall, and in countries where nursing homes are widely used, the difference was very substantial. In secondary analyses, the aggregated estimates show very low IFR estimates for younger age groups.

Early estimates of case fatality rate (CFR, ratio of deaths divided by *documented* infections) in the elderly were very high and they played an instrumental role in disseminating both fear and alacrity in dealing with this serious pandemic. Early estimates of CFR from China (58) described CFR of 8% in the age group 70-79 and 14.8% in those ≥80 years. Extremely high CFR estimates were also reported initially from Italy (59) and New York (60). However, the number of infected individuals was much larger than the documented cases (61). Therefore, IFR is much lower than CFR. We are aware of three previous evaluations of age-stratified IFR estimates that combine seroprevalence data with age-specific COVID-19 mortality statistics (4, 5, 62). Whereas the current report is the only one to use the term “mixed-design” synthesis, also previous syntheses of IFRs using seroprevalence data are, by necessity, mixed-design, although decisions and calculations may not be as transparently disclosed. This work represents the only effort to date to synthesize data on age-stratified IFR estimates that uses a highly detailed prespecified and registered protocol, with addition of detailed justifications whenever further decisions had to be made. An exception is the secondary (non-prespecified) analysis of IFR in younger age groups; however, we argue that the eligibility criteria (consideration of large studies with >1000 elderly individuals) are not likely to have introduced bias specifically for the IFR estimates in younger age groups.

Levin *et al* (4) is the basis for the US CDC pandemic planning scenarios (63). Levin *et al* report IFR 4.6% at age 75, and 15% at age 85 (4) without separating nursing home deaths (thus referring to all elderly). The assessment was based on relatively sparse data for these age groups and was limited to advanced economies. The authors counted deaths four weeks after the midpoint of the seroprevalence sampling period, which is the longest among the evaluations, with the argument that there is large potential reporting lag (although available mortality statistics are commonly updated retrospectively for the date of death). Also, almost all included studies came from hard-hit locations, where IFR may be substantially higher (3). Selection bias for studies with lower seroprevalence and/or higher death counts (6) may explain why their estimates for middle-aged and elderly are substantially higher than ours.

O’Driscoll *et al* (5) modeled 22 seroprevalence studies, and carefully comment how outbreaks in nursing homes can drive overall population IFRs. For young and middle-aged groups, their estimates largely agree with those presented here. Their estimates for elderly, which are reflecting the community-dwelling, are still higher than ours. For ages ≥65 years, their model uses data derived from one location (England) on deaths that did not occur in nursing homes and is validated against other locations with such statistics. This may overestimate the community-dwelling proportion, since deaths of nursing home residents occurring in hospitals are counted in the England community estimates. Conversely our evaluation adds granularity by using deaths in nursing home residents from many countries, and by using seroprevalence estimates from 25 serosurveys with many elderly individuals.

The Imperial College COVID-19 response team (62) presents much higher IFR estimates for elderly overall. They use a very narrowly selected subset of 10 studies in 9 countries, five of which had sampled >1000 elderly people. Their selection criteria required >100 deaths in the location at the seroprevalence study midpoint, which skews the sample towards heavily-hit areas and higher IFRs (6).

Some published studies also present IFRs in elderly people for single locations based on seroprevalence data, but these are unavoidably location-limited (see Appendix text).

For persons 0-19 years, the median IFR was one death per 76,900 persons with COVID-19 infection, followed by estimates of 1:11,300 in ages 20-29, 1:4800 in ages 30-39, and 1:2400 in ages 40-49. The Imperial College study (62) has ∼10 times higher estimates for persons 0-19 years and ∼3 times higher for persons 20-29 years old; otherwise estimates in age groups <50 years are fairly consistent across previous (4, 5) and current analyses despite methodological differences.

Substantial true heterogeneity is very much expected since IFR is situation- and population-dependent. Both the age distribution and other characteristics of people within the elderly stratum vary between different countries. E.g., obesity is a major risk factor for poor outcome with COVID-19 infection and prevalence of obesity is only 4% in India versus 20-36% in high-income countries analyzed here. Besides differences in risk factor characteristics, documentation criteria for coding COVID-19 deaths may have varied non-trivially across countries. Under- and over-counting of COVID-19 deaths may have occurred even in countries with advanced health systems.

The observed differences in IFR between community-dwelling elderly and elderly overall is consistent with previous findings that beyond age, comorbid conditions and frailty are associated with higher COVID-19 mortality (e.g., (10)). Given that nursing home residents account for many COVID-19 deaths (64), a location’s overall IFR across all ages is largely dependent on how nursing homes were afflicted (5). Spread in nursing homes was disproportionately high in the first wave (8). IFR in nursing home residents can be much higher than the IFR estimates we obtained for community-dwelling elderly. Seroprevalence studies of long-term care populations in Spain (Madrid), northern Italy, UK and Brazil in early phases of the epidemic found prevalence of 55%, 41%, 33%, and 11.5%, respectively (65-69), i.e. several fold higher than the prevalence in the general community populations in these locations. Under-estimation of infections due to seroreversion may also be prominent in such studies (69). Large diversity in seroprevalence can exist between facilities, e.g. in the Brazil study (68) the seroprevalence was 100% and 76%, respectively in 2 nursing homes that had outbreaks and 0% or close to 0% in the other 13. The IFR in that study was 25% and this may be a reasonable estimate for nursing homes with rather debilitated frail populations, as corroborated also by other investigators (11). IFR may be much lower in facilities where residents are in overall good health and plan to spend many years of their life there; but extremely high in facilities that offer primarily palliative care.

The share of nursing home deaths decreased markedly over time (64) in most high-income countries with some exceptions (e.g. Australia). This change may be reflected in a much lower IFR among the elderly and the entire population after the first wave. Improved treatments (e.g. dexamethasone), and less use of harmful treatments (e.g. hydroxychloroquine, improper mechanical ventilation) may also have decreased IFR substantially in late 2020 and in 2021 (70, 71). Other investigators have estimated in 2020 a global IFR of only 0.11% in the absence of effects of new variants and vaccinations (72). Vaccines that are more effective in protecting against death rather than infection are also expected to have decreased the IFR in 2021. New variants becoming dominant in 2021 may also be associated with further lower IFR. E.g., in countries with extensive testing such as the UK, when the delta variant spread widely even CFR remained ∼0.3% (73). Preliminary data on the omicron variant in late 2021 suggest that it may be associated with even lower severity (74).

Our analysis has several limitations. First, seroprevalence estimates among elderly reported by the included studies could over- or underestimate the proportion infected. We explored adjusted estimates accounting for 1-10% relative seroreversion per month; however, higher seroreversion is likely (21, 22, 32). Higher seroreversion will affect more prominently studies carried out later in the pandemic. Also, the current estimates do not fully account for the unknown share of people who may have tackled the infection without generating detectable serum/plasma antibodies (e.g., by mucosal, innate, or cellular [T cell] immune mechanisms) (75-79). Sensitivity estimates for antibody assays typically use positive controls from symptomatic individuals with clinically manifest infection; sensitivity may be lower for asymptomatic infections. All seroprevalence studies may have substantial residual biases despite whatever adjustments, as is discussed in detail by one of us (JPAI) elsewhere, in terms of potential biases of different types of designs and sampling frames regarding representativeness against the general population (6). Even well-designed general population studies may specifically fail to reach and recruit highly vulnerable populations, e.g. disadvantaged groups, immigrants, homeless, and other people at high exposure risks and poor health. For studies carried out in the US, we prespecified an eligibility criterion to adjust for race/ethnicity, which we believe acts to mitigate related biases. Definitions and usage of race and ethnicity variables were expected to be too heterogeneous outside the US to introduce a common rule, which would exclude virtually all studies from non-US locations (as can be seen in Table 1, only one other study adjusts for ethnicity, one adjusts for nationality, and even other factors associated with socioeconomic position are scarce). The study by Paulino-Ramirez et al (41), in the Dominican Republic, specifically targeted “communities identified as emerging hotspots for SARS-CoV-2” for antibody testing, and should be mentioned specifically to have a risk of overestimating seroprevalence. This especially has relevance for generalizability of IFR to other middle-income countries, but has small bearing on the median IFR in the sample overall.

Second, the number of deaths may be biased for various reasons (3) leading to potential under- or over-counting. Among the countries included in this synthesis, others have pointed out concerns of underreporting of deaths specifically for India (e.g., (80)). Indeed, IFR estimates in India need to be seen with extra caution. A study in Madurai in south India found an IFR of 0.043% among people >15 years old, but estimated that given the age distribution the IFR should have been 9 times larger to match other countries (81). This could reflect under-reporting of deaths, lower rates of age-adjusted death risk in India, or both (82). For example, compared with USA and European countries, India has much lower proportions of obesity, diabetes, smoking, heart disease, medically maintained patients with terminal cancer, and medically immunosuppressed patients. These are all strong risk factors for fatality from COVID-19. Also of note, excess deaths in India in 2020-2021 may be much higher than the reported COVID-19 toll (83). However, excess deaths in a single year are notoriously difficult to calculate (especially in a country with suboptimal death registration) and reflect the composite of direct deaths due to COVID-19, indirect effects of the pandemic, direct and indirect effects of the measures taken and multiple other year- and country-specific causes (84, 85). Excess deaths should not be used to calculate IFR.

Of note, our estimates of IFRs for non-elderly young strata in India are not very dissimilar to those of high-income countries, while the divergence is much stronger in the elderly population. This may be further explained by the fact that among people >70 years old, the proportion of those who are >85 years old is only 9.5% in India, while it is much higher in high income countries (e.g. Spain 23.7%, Italy 21.1%). Moreover, the difference between India and high-income countries in prevalence of major comorbidities such as obesity that increase the risk of COVID-19 mortality is most prominent in the most elderly; in younger generations in India, the influence of western lifestyle is becoming more pervasive (86, 87). Therefore, the IFR in people >70 years old in India may well be extremely lower than the respective figure in high-income countries. The same may apply also to other middle-income countries and also low-income countries.

To match the date for seroprevalence sampling (i.e., seroconversion) with cumulative deaths is an exercise with assumptions. Our sensitivity analysis that extended with one week the cutoff for counting deaths showed a negligible change in the median IFR calculation. Most studies included in our analysis had been performed during periods at or after the end of the first wave. Some studies performed sampling for several months, which introduces further uncertainty. However, typically the sampling covered periods with few fatalities, why the uncertainty about corresponding death counts and infection rates would arguably be small.

Third, we acknowledge the risk of bias in seroprevalence studies, mortality statistics, and even population statistics. However, assessments of risk of bias are far from straightforward, as illustrated by the discrepant assessments of these seroprevalence studies by other teams (6). It should be expected that estimates cited here have considerable uncertainty that is not accounted for in the presented 95% CIs, which were based solely on seroprevalence study sampling uncertainty (with potential adjustments). Others, e.g., Campbell and Gustafson (88), have proposed models that aim to take into account further sources of uncertainty. Overall, the strength of evidence regarding COVID-19 mortality has markedly improved since the early days of the pandemic, although some biases are still affecting currently available studies.

Fourth, even among high-income countries, our set of eligible surveys tends to include mostly data from countries with higher death rates, thus possibly also higher IFR. More prominently, our analysis includes limited data from Asia and no data from Africa. Consideration of age strata diminishes this representativeness bias, but cannot eliminate it. E.g., most countries not represented in the available data may have a shift towards lower ages within the stratum of the elderly. This translates to lower IFR. Moreover, with the exception of India, all countries analyzed here have population prevalence of obesity 1.5-3-fold higher than the global prevalence (13%); other major risk factors for poor COVID-19 outcome such as smoking history, diabetes, cardiovascular disease, and immunosuppression (9) are also far more common in the high-income countries included in our analysis than the global average. Global IFR may thus be substantially lower in both the elderly and the lower age strata than estimates presented herein.

Fifth, as for previous syntheses of IFR based on seroprevalence data, many complementary pieces of information were needed beyond the systematic search for seroprevalence studies. Some of the decisions made in this process, e.g., the eligibility criterion of >1000 participants, could be described as arbitrary. This rule was introduced for feasibility and validity, since over a thousand of seroprevalence studies were available, the majority of which would be largely uninformative (given the huge uncertainty) even for the overall population, let alone the rarified subgroup of elderly persons. As we mention elsewhere, it tends to prefer studies done in populations with old age pyramids, and this may thus over-represent populations with many frail individuals who survive into late age with many comorbidities. Importantly, none of these rules were expected to introduce other overt biases, and they were set *a priori*.

This overview synthesis finds a consistently much lower IFR of COVID-19 in community-dwelling elderly than in elderly overall, a difference which is substantial in countries where nursing homes are an established form of residency. Very low IFR estimates were confirmed in younger groups (<50). For middle-aged groups and elderly, estimates were lower than in some previous influential work with biased methodological choices (4, 62), but in agreement with other work (5). The estimates presented here may serve as one of several key pieces of information underlying public health policy decisions. With better management and better preventive measures hopefully IFR has already decreased further.

## Data Availability

The protocol, data, and code used for this analysis will be made available at the Open Science Framework upon publication.

https://osf.io/47cgb

## ACKNOWLEDGMENTS

We thank sincerely Eva Heras Muxella, Cristina Royo-Cebrecos (Andorra), Sereina Herzog (Belgium), Ilse Peeters and Catharina Vernemmen at Sciensano, the Belgian Institute for Health (Belgium), Zoltán Vokó (Hungary), Peter Coyle, Laith Abu-Raddad (Qatar), for personal communication of seroprevalence and/or COVID-19 mortality statistics; and Giridhara D Babu, Rajesh Sundaresan, Siva Athreya (India), N. Ahmad Aziz, Monique M.B. Breteler (Germany), Public Health Scotland (Scotland), Office for National Statistics, Infection Survey Analysis (UK), Cesar Victora, Fernando Hartwig, Luis Vidaletti (Brazil) for personal communication to clarify non-eligibility. We thank the dedicated members of the SeroTracker and Worldometer teams as well as Wikipedia authors for their efforts to gather information on seroprevalence studies and COVID-19 mortality statistics.

## DATA SHARING STATEMENT

The protocol, data, and code used for this analysis will be made available at the Open Science Framework upon publication: https://osf.io/47cgb.

## TRANSPARENCY STATEMENT

The lead author affirms that the manuscript is an honest, accurate, and transparent account of the study being reported; that no important aspects of the study have been omitted; and that any discrepancies from the study as originally planned have been explained.

## LICENSE STATEMENT

The Corresponding Author has the right to grant on behalf of all authors and does grant on behalf of all authors, a worldwide licence to the Publishers and its licensees in perpetuity, in all forms, formats and media (whether known now or created in the future), to i) publish, reproduce, distribute, display and store the Contribution, ii) translate the Contribution into other languages, create adaptations, reprints, include within collections and create summaries, extracts and/or, abstracts of the Contribution, iii) create any other derivative work(s) based on the Contribution, iv) to exploit all subsidiary rights in the Contribution, v) the inclusion of electronic links from the Contribution to third party material where-ever it may be located; and, vi) licence any third party to do any or all of the above.

## CONTRIBUTOR STATEMENT

The corresponding author attests that both listed authors meet authorship criteria and that no others meeting the criteria have been omitted.

Cathrine Axfors: Conceptualization, Methodology, Software, Formal analysis, Investigation, Data curation, Writing - original draft, Visualization, Project administration.

John P A Ioannidis: Conceptualization, Methodology, Formal analysis, Investigation, Data curation, Writing - review & editing, Visualization, Supervision.

