## Appendix for "Infection fatality rate of COVID-19 in community-dwelling populations with emphasis on the elderly: An overview"

Cathrine Axfors[1], John P A Ioannidis[1,2]
[1]Meta-Research Innovation Center at Stanford (METRICS), Stanford University, Stanford, California, USA
[2]Departments of Medicine, of Epidemiology and Population Health, of Biomedical Data Science, and of Statistics, Stanford University, Stanford, California, USA

Correspondence to: John P A Ioannidis, Stanford Prevention Research Center, Medical School Office Building, Room X306, 1265 Welch Road, Stanford CA 94305, USA

1. **Appendix Figure 1.** Flowchart of the search and selection process.
2. **Appendix Figure 2**. Flowchart of the search and selection process, sensitivity analysis: Including explicitly national-level general population studies without high risk of bias and with at least 500 participants aged ≥70 years.
3. **Appendix Figure 3.** Age-specific infection fatality rates (IFRs) in this work and some previously published overviews (log-scale and normal scale).
4. **Appendix Table 1**. Amendments to protocol
5. **Appendix Table 2**. Sources for COVID-19 mortality statistics and population statistics.
6. **Appendix Table 3a.** Reports not included, which sampled or potentially sampled ≥1000 participants ≥70 years of age.
7. **Appendix Table 3b.** Reports not included, sensitivity analysis: Including explicitly national-level general population studies without high risk of bias and with at least 500 participants aged ≥70 years.
8. **Appendix Table 4.** Seroprevalence estimates corrected for test performance using the Gladen-Rogan formula.
9. **Appendix Table 5.** Uncorrected and seroreversion-corrected infection fatality rate in community-dwelling elderly
10. **Appendix Table 6**. Sensitivity analysis with a later cutoff for cumulative COVID-19 mortality (study midpoint plus two weeks instead of plus one week).
11. **Appendix text**

**Appendix Figure 1.** Flowchart of the search and selection process.

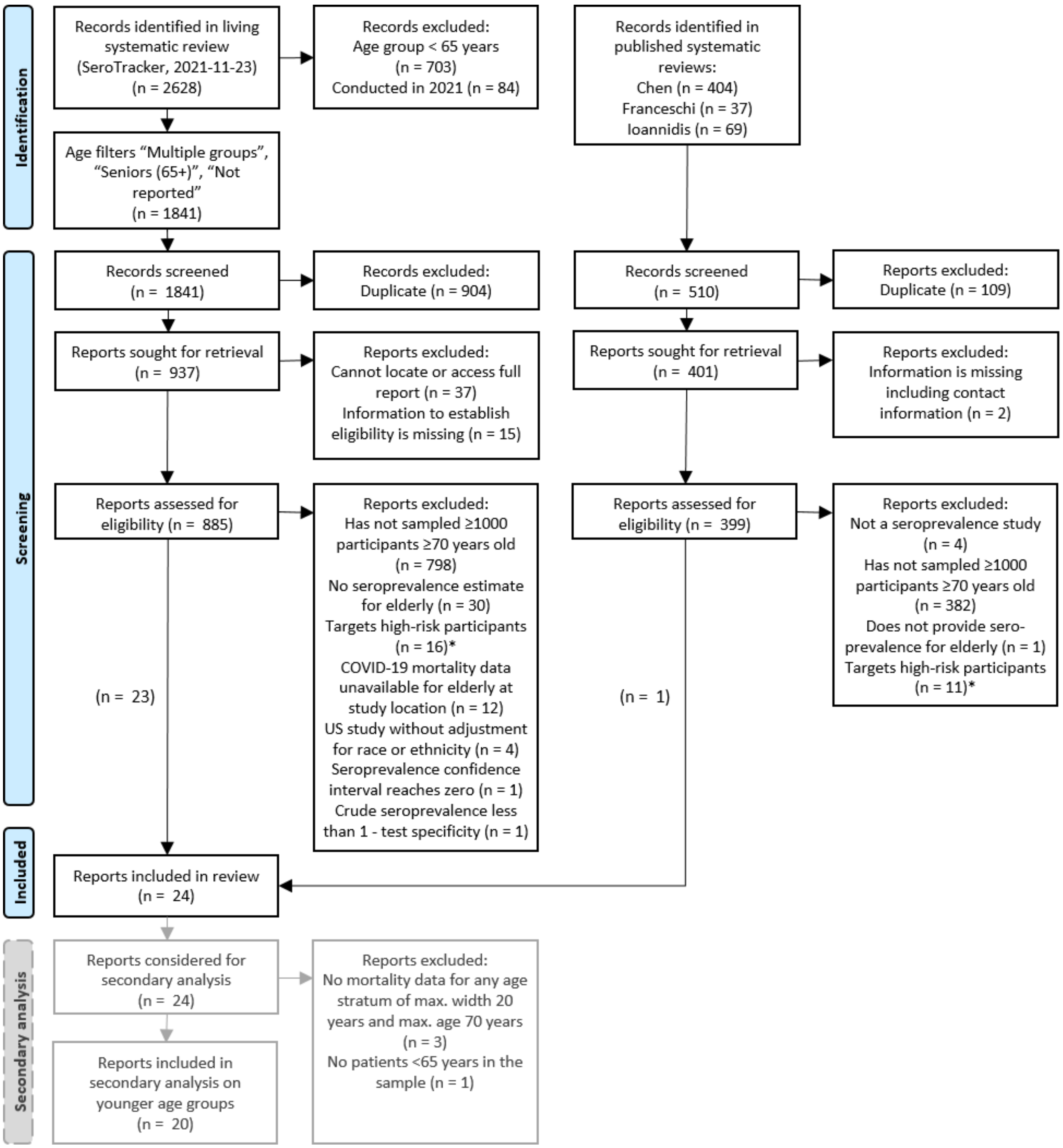

Systematic reviews used as sources: SeroTracker (7); Chen et al (12), Franceschi et al (11), Ioannidis (13). SeroTracker is a living systematic review with daily screening of published articles (MEDLINE, Embase, Web of Science, Cochrane), preprints (medRxiv, bioRxiv), government reports, and news articles. Data on identified seroprevalence studies are available for downloading at www.serotracker.com including study characteristics such as sampled age groups.

*High-risk participants: healthcare workers, essential workers, contacts to COVID-19 patients, or residual patient samples where COVID-19-related visits were not excluded. In the final sample (24 reports for main analysis; 20 reports for secondary analysis), one report presented data on two eligible surveys.

**Appendix Figure 2.** Flowchart of the search and selection process, sensitivity analysis: Including explicitly national-level general population studies without high risk of bias and with at least 500 participants aged ≥70 years.

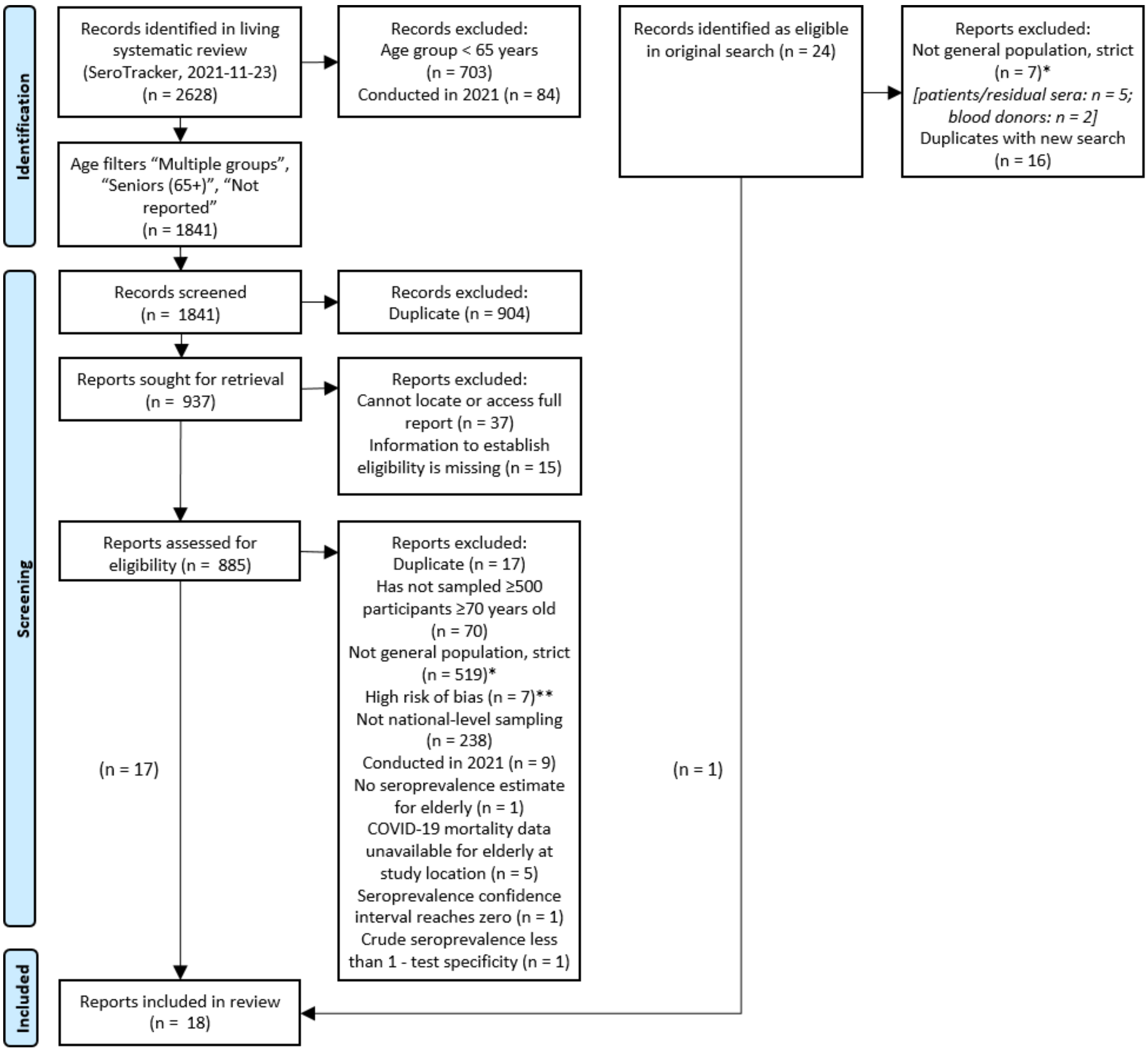

*A stricter criterion was used for general population samples in this sensitivity analysis than in the main analysis, based on SeroTracker categories of "Household and community samples" and "Multiple general populations". ** Risk of bias according to assessments by the SeroTracker team.

In the final sample (18 reports), one presented data on two eligible surveys.

**Appendix Figure 3**. Age-specific infection fatality rates (IFRs) in this work and some previously published overviews (log-scale and normal scale).

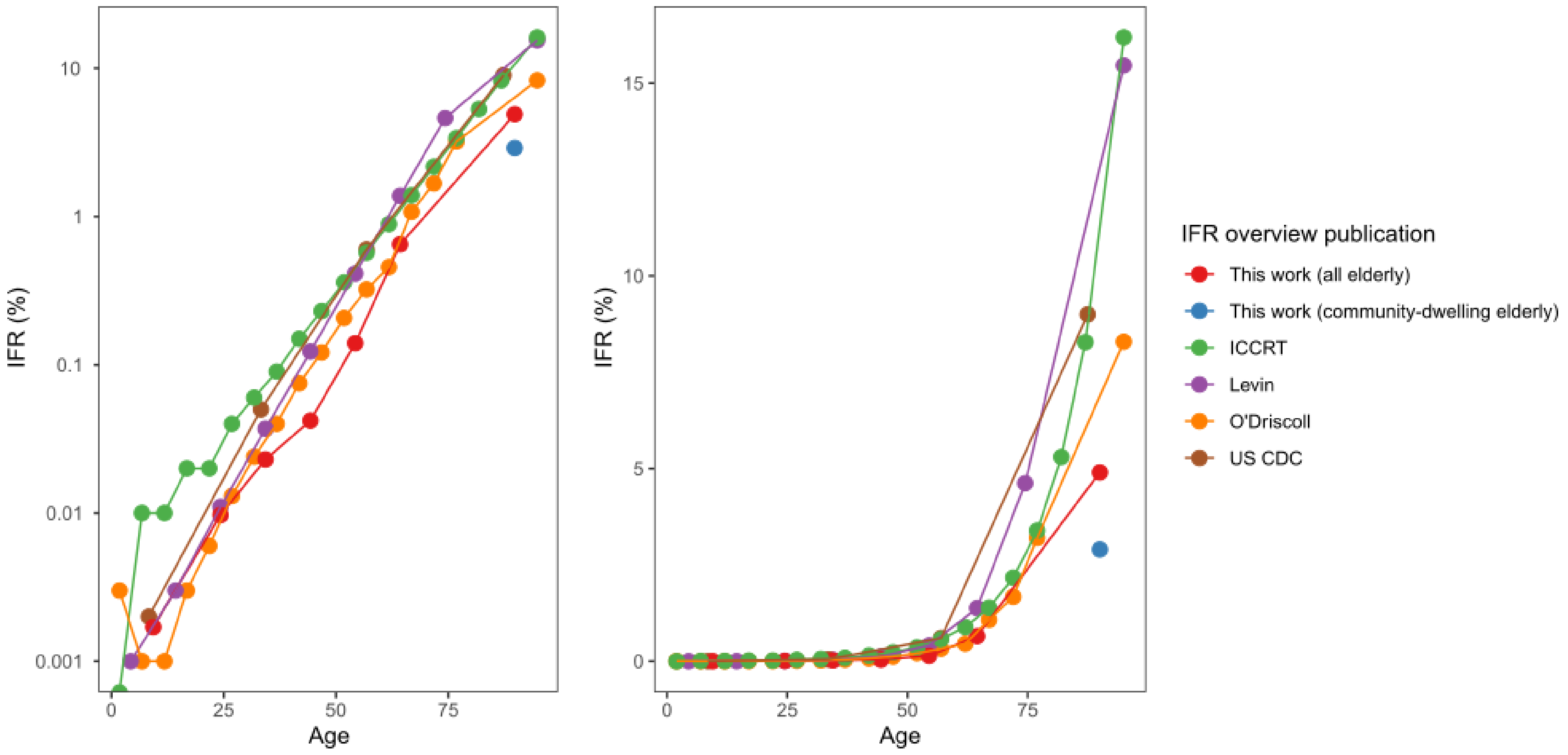

The eldest groups, which do not have an upper age bound, are placed at the midpoint of their lower bound and 110. CDC: Centers for Disease Control and Prevention. ICCRT: Imperial College COVID-19 response team. Estimates for O'Driscoll regarding age brackets 65 years and older refer to the community-dwelling, while other estimates outside this work refer to all elderly.

**Appendix Table 1**. Amendments to protocol.*

| **Manuscript section** | **Amendment** | **Rationale** |
|---|---|---|
| Eligibility criteria | Added eligibility criterion: excluding seroprevalence studies that sampled participants after Dec 31, 2020. | IFR may be substantially affected (mostly shifting downwards) by vaccination (especially with prioritization among the most aged and frail individuals), better treatment, and new variants emerging and becoming dominant in 2021. We located only one study with sampling in 2021 (Office of National Statistics, United Kingdom, February 2021). |
| Eligibility criteria | Added eligibility criterion: excluding seroprevalence studies where crude seroprevalence is less than 1- test specificity and/or the 95% confidence interval of the seroprevalence goes to 0%. | Following comments from peer-reviewers, since the seroprevalence estimate would be extremely uncertain. |
| Eligibility criteria | Added clarification: For studies with several sampled subregions of a country we applied the criterion of ≥1000 sampled individuals ≥70 years for each subregion, unless (1) the sampling locations were dispersed across the country so as to form a reasonable representation of the entire country, in which case the country was considered the location; or (2) an aggregate seroprevalence estimate was available *and* age-stratified COVID-19 mortality statistics and population statistics were available for each subregion, in which case the regions combined were considered the location. | Clarification was necessary for this eligibility criterion in the case of studies with several sampled subregions. As far as possible, we strived to combine subregional sampling with the respective COVID-19 mortality and population statistics. |
| Data extraction | Added clarification: We extracted the number of deaths for the primary date from official reports or Worldometer (or, if the location was not present on Worldometer, Wikipedia), whichever presented the higher number. For official reports, we preferred the latest updated dataset of deaths by date of occurrence (not date of reporting) if available, otherwise situational reports. | Minor discrepancies were observed between sources, possibly attributable to retrospectively reported deaths, why the higher number was chosen. Latest available datasets were preferred to situational reports because of the addition of retrospectively reported deaths. |
| Data extraction | If the number of deaths occurring in nursing homes (i.e., excluding deaths occurring in hospitals) was the only available estimate, we calculated the total number of deaths in nursing home residents with a correction (by multiplying with the median of available ratios of deaths in nursing homes to deaths of nursing home residents in the International Long Term Care Policy Network report of October 14 of countries in the same continent). | In order to not count the deaths of nursing home residents occurring in hospital as deaths in community-dwelling elderly. The correction was applied to one study (Hughes). |
| Sensitivity analyses | The seroreversion correction is calculated with a $X^m$-fold correction to the IFR, not $(1/X)^m$-fold. | Correction of typo in protocol. |
| Sensitivity analyses | Added clarification: The peak of the first epidemic wave was defined as one week before the date with the highest rolling average 7-day mortality (according to Worldometer, situational reports, or Wikipedia). The first epidemic wave was defined to end by the date with the lowest 7-day average of daily deaths since the beginning of the epidemic. If two or more dates were tied for peak values, we chose the date corresponding to the midpoint between the first and last one. | This definition had been missed in the protocol. |
| Sensitivity analyses | Added sensitivity analysis: focusing only on explicitly general population samples based on the SeroTracker categories of "Household and community samples" and "Multiple general populations" and thus excluding for example patient cohorts, insurance applicants, blood donors, and workers, | Following comments from peer-reviewers |

| | | |
|---|---|---|
| Sensitivity analyses | Added sensitivity analysis: including only explicitly national-level general population studies without high risk of bias and with at least 500 participants aged ≥70 years. Explicitly general population samples were based on the SeroTracker categories of "Household and community samples" and "Multiple general populations" and thus excluding for example patient cohorts, insurance applicants, blood donors, and workers. We applied the risk of bias assessments reported by the SeroTracker team (based on the Joanna Briggs Institute Critical Appraisal Tool for Prevalence Studies). | Following comments from peer-reviewers |
| Sensitivity analyses | Added sensitivity analysis: including only studies conducted in high-income countries | Following comments from peer-reviewers |
| Sensitivity analyses | Added sensitivity analysis: excluding studies where the selected time point with highest seroprevalence was not the latest available (seroprevalence had declined in the latest timepoint). | Following comments from peer-reviewers |
| Data Synthesis and Analysis | We added a calculation of 95% CIs of IFRs based on extracted 95% CIs from seroprevalence estimates. Primarily, 95% confidence intervals are direct extractions from the seroprevalence studies. For studies that did not report such intervals, we complemented with a calculation using the number of sampled and seropositive elderly individuals (Clopper Pearson interval calculation).<br>For those that provided adjusted estimates for age brackets (e.g., 70-79, 80-89, and 90+), we combined estimates for each study using a fixed effects inverse variance meta-analysis (of arcsine transformed proportions) to obtain 95% CIs.<br>No further factors were introduced in the calculation beyond the adjustments made by seroprevalence study authors (except adjusting estimates for test performance using the Gladen-Rogan formula where applicable, see below). | In order to illustrate in Figure 1 the uncertainty of IFR estimates arising from seroprevalence sampling as reported by seroprevalence study authors, and to calculate the heterogeneity indicator I-squared.<br>The 95% CIs of IFRs do not take into account other sources of uncertainty than those adjusted by the seroprevalence authors, and should be interpreted as very conservative. |
| Calculated Data Variables | Added clarification: For studies that provided adjusted seroprevalence estimates for age brackets (e.g., 70-79, 80-89, and 90+) we calculated the number of infected individuals for each age bracket by multiplying with the corresponding population size. (For community-dwelling elderly, we first subtracted the nursing home population, assuming that the proportion of nursing home residents was equal across the age brackets). Then, we summarized the number of infected individuals in the age brackets to the number of infected individuals for (community-dwelling) elderly. | In order to be able to use adjusted seroprevalence estimates that were provided for age brackets that together formed an eligible elderly stratum. |
| Calculated Data Variables | For Qatar, and for the Dominican Republic, we could not retrieve information on the proportion of deaths accounted by nursing home residents, and we imputed a value of 10%. Imputation for these countries were not prespecified. | Qatar has a small number of nursing home residents relative to the population, although the number has increased in recent years. For the Dominican Republic, we imputed the same number prespecified for India. |
| Calculated Data Variables | We applied a non-prespecified correction for studies that excluded persons with diagnosed COVID-19 from participating in their sample, primarily using study authors' corrections (e.g., PCR tests) or adding the number of identified COVID-19 cases in community-dwelling elderly for the location until the seroprevalence study midpoint. | Added in order to represent correctly the number of infected persons. |

| | | |
|---|---|---|
| | For Gudbjartsson et al, we used the numbers based on seroprevalence and detection of ongoing infections.<br><br>For Kalish et al, we calculated the number of cases in community-dwelling elderly up to June 20 by retrieving the cumulative number of cases in elderly (≥65) from the COVID-19 Data Tracker at the US CDC (455,585) and subtracting the cumulative number of cases in nursing homes from KFF (240,138), resulting in 215,447 cases. This number was added to the number of infected community-dwelling elderly. | |
| Calculated Data Variables | Added calculation: Whenever no adjustment was made for test performance, we adjusted the estimates for test performance using the Gladen-Rogan formula (27). | Following comments from peer-reviewers |

* Protocol, Open Science Framework: https://osf.io/47cgb

**Appendix Table 2**. Sources for COVID-19 mortality statistics and population statistics.

| |
|---|
| **Andorra (Study: Royo-Cebrecos)** |
| Seroprevalence information: Seroprevalence study |
| Age distribution of COVID-19 deaths: Personal communication E. Heras Muxella and C. Royo-Cebrecos |
| Total number of deaths for the primary date: Personal communication E. Heras Muxella and C. Royo-Cebrecos |
| COVID-19 deaths in nursing home residents: Personal communication E. Heras Muxella and C. Royo-Cebrecos |
| Peak 7-day average of COVID-19 mortality: Worldometers.info |
| Population overall and in elderly: https://www.estadistica.ad/serveiestudis/web/banc_dades4.asp?lang=4&codi_tema=2&codi_divisio=2783&codi_subtemes=8 |
| Population in nursing homes: Personal communication E. Heras Muxella and C. Royo-Cebrecos |
| Proportion of elderly among nursing home residents: Imputed (average of available values) |
| Comments: Mortality peak is the midpoint of April 1, 2020, and April 16, 2020. |
| **Belgium (Study: Herzog)** |
| Seroprevalence information: Seroprevalence study and personal communication S. Herzog |
| Age distribution of COVID-19 deaths: Personal communication I. Peeters, Sciensano, the Belgian Institute for Health (For younger age groups: Riffe, T., Acosta, E., the COVerAGE-DB team, Data Resource Profile: COVerAGE-DB: a global demographic database of COVID-19 cases and deaths, International Journal of Epidemiology, Volume 50, Issue 2, April 2021, Pages 390–390f, https://doi.org/10.1093/ije/dyab027. Accessed online as 10.17605/OSF.IO/MPWJQ.) |
| Total number of deaths for the primary date: Personal communication I. Peeters, Sciensano, the Belgian Institute for Health (number as of April 1, 2020) |
| COVID-19 deaths in nursing home residents: Situational report (COVID-19 – BULLETIN EPIDEMIOLOGIQUE DU 10 AVRIL 2020, Sciensano) retrieved from https://covid-19.sciensano.be/fr/covid-19-situation-epidemiologique |
| Peak 7-day average of COVID-19 mortality: Worldometers.info |
| Population overall and in elderly (Jan 1, 2020): Dataset (Population de la Belgique par âge, au 1er janvier) retrieved from https://www.plan.be/databases/data-35-fr-perspectives_de_population_2020_2070 |
| Population in nursing homes: Personal communication I. Peeters, Sciensano, the Belgian Institute for Health |
| Proportion of elderly among nursing home residents: Imputed (average of available values) |
| Comments: Only using Collection period 1 (Mar 30-Apr 5) while there are 4 more, ending July 4 (none of the others have >1000 elderly). |
| **Canada (Study: Saeed, Canadian Blood Services)** |
| Seroprevalence information: Seroprevalence study |
| Age distribution of COVID-19 deaths: Riffe, T., Acosta, E., the COVerAGE-DB team, Data Resource Profile: COVerAGE-DB: a global demographic database of COVID-19 cases and deaths, International Journal of Epidemiology, Volume 50, Issue 2, April 2021, Pages 390–390f, https://doi.org/10.1093/ije/dyab027. Accessed online as 10.17605/OSF.IO/MPWJQ. |
| Total number of deaths for the primary date: Riffe, T., Acosta, E., the COVerAGE-DB team, Data Resource Profile: COVerAGE-DB: a global demographic database of COVID-19 cases and deaths, International Journal of Epidemiology, Volume 50, Issue 2, April 2021, Pages 390–390f, https://doi.org/10.1093/ije/dyab027. Accessed online as 10.17605/OSF.IO/MPWJQ. |
| COVID-19 deaths in nursing home residents: Comas-Herrera A, Zalakaín J, Litwin C, Hsu AT, Lemmon E, Henderson D and Fernández J-L (2020) Mortality associated with COVID-19 outbreaks in care homes: early international evidence. Article in LTCcovid.org, International Long-Term Care Policy Network, CPEC-LSE, 26 June 2020. |
| Peak 7-day average of COVID-19 mortality: Worldometers.info |
| Population overall and elderly: StatCan Census 2016, dataset retrieved from https://www12.statcan.gc.ca/census-recensement/2016/dp-pd/index-eng.cfm |
| Population in nursing homes: StatCan Census 2016, dataset retrieved from https://www12.statcan.gc.ca/census-recensement/2016/dp-pd/index-eng.cfm |
| Proportion of elderly among nursing home residents: StatCan Census 2016, dataset retrieved from https://www12.statcan.gc.ca/census-recensement/2016/dp-pd/index-eng.cfm |
| **Canada (Study: Tang / Ab-C Study)** |
| Seroprevalence information: Seroprevalence study |
| Age distribution of COVID-19 deaths: Seroprevalence study |
| Total number of deaths for the primary date: Riffe, T., Acosta, E., the COVerAGE-DB team, Data Resource Profile: COVerAGE-DB: a global demographic database of COVID-19 cases and deaths, International Journal of Epidemiology, Volume 50, Issue 2, April 2021, Pages 390–390f, https://doi.org/10.1093/ije/dyab027. Accessed online as 10.17605/OSF.IO/MPWJQ. |

| |
|---|
| COVID-19 deaths in nursing home residents: Seroprevalence study |
| Peak 7-day average of COVID-19 mortality: Worldometers.info |
| Population overall: Seroprevalence study |
| Population in elderly: Seroprevalence study |
| Population in nursing homes: Seroprevalence study |
| Proportion of elderly among nursing home residents: Seroprevalence study |
| Comments: Definition of nursing home in this study includes mixed care and retirement homes.<br>Confidence intervals for adjusted seroprevalence were calculated using the inferred number of adjusted seropositive individuals.<br>Note the alternative definition of "possible" positives depending on which controls are used, here: "Definite or possible". |
| **Canada [Ontario] (Study: Public Health Ontario)** |
| Seroprevalence information: Personal communication Public Health Ontario Communications |
| Age distribution of COVID-19 deaths: Enhanced Epidemiological Summary (COVID-19 Case Fatality, Case Identification, and Attack Rates in Ontario), May 17, retrieved from https://www.publichealthontario.ca/en/data-and-analysis/infectious-disease/covid-19-data-surveillance/weekly-epi-summary-archive |
| Total number of deaths for the primary date: Wikipedia |
| COVID-19 deaths in nursing home residents: Dataset at https://data.ontario.ca/en/dataset/long-term-care-home-covid-19-data |
| Peak 7-day average of COVID-19 mortality: Dataset at https://covid-19.ontario.ca/data (accessed April 30, 2021) |
| Population overall and in elderly: StatCan Census 2016, dataset at https://www12.statcan.gc.ca/census-recensement/2016/dp-pd/index-eng.cfm |
| Population in nursing homes: StatCan Census 2016, dataset at https://www12.statcan.gc.ca/census-recensement/2016/dp-pd/index-eng.cfm |
| Proportion of elderly among nursing home residents: StatCan Census 2016, dataset at https://www12.statcan.gc.ca/census-recensement/2016/dp-pd/index-eng.cfm |
| Comments: 100% of the nursing home deaths reported were in elderly 70+. |
| **Canada [Alberta] (Study: Charlton)** |
| Seroprevalence information: Seroprevalence study |
| Age distribution of COVID-19 deaths: Riffe, T., Acosta, E., the COVerAGE-DB team, Data Resource Profile: COVerAGE-DB: a global demographic database of COVID-19 cases and deaths, International Journal of Epidemiology, Volume 50, Issue 2, April 2021, Pages 390–390f, https://doi.org/10.1093/ije/dyab027. Accessed online as 10.17605/OSF.IO/MPWJQ. |
| Total number of deaths for the primary date: Riffe, T., Acosta, E., the COVerAGE-DB team, Data Resource Profile: COVerAGE-DB: a global demographic database of COVID-19 cases and deaths, International Journal of Epidemiology, Volume 50, Issue 2, April 2021, Pages 390–390f, https://doi.org/10.1093/ije/dyab027. Accessed online as 10.17605/OSF.IO/MPWJQ. |
| COVID-19 deaths in nursing home residents: Canadian Institute for Health Information. The Impact of COVID-19 on Long-Term Care in Canada: Focus on the First 6 Months. Ottawa, ON: CIHI; 2021. |
| Peak 7-day average of COVID-19 mortality: COVID-19 Alberta Statistics. Summary data starting March 6, 2020. Government of Alberta. Retrieved from https://www.alberta.ca/stats/covid-19-alberta-statistics.htm#data-export |
| Population overall and in elderly: StatCan Census 2016, dataset at https://www12.statcan.gc.ca/census-recensement/2016/dp-pd/index-eng.cfm |
| Population in nursing homes: StatCan Census 2016, dataset at https://www12.statcan.gc.ca/census-recensement/2016/dp-pd/index-eng.cfm |
| Proportion of elderly among nursing home residents: StatCan Census 2016, dataset at https://www12.statcan.gc.ca/census-recensement/2016/dp-pd/index-eng.cfm |
| Comments: For COVID-19 deaths in nursing home residents, the number for Alberta, Canada, was unavailable historically. We therefore extrapolate from the Canada proportion of COVID-19 deaths in nursing home residents among all deaths (up to February 15, 2021): 67%, see reference listed above. |
| **Denmark [3 regions] (Study: Pedersen)** |
| Seroprevalence information: Seroprevalence study |
| Age distribution of COVID-19 deaths: Seroprevalence study |
| Total number of deaths for the primary date: Dataset at https://covid19.ssi.dk/overvagningsdata/download-fil-med-overvaagningdata [Deaths_over_time.csv] (Accessed February 17, 2021) |
| COVID-19 deaths in nursing home residents: Comas-Herrera A, Zalakaín J, Litwin C, Hsu AT, Lemmon E, Henderson D and Fernández J-L (2020) Mortality associated with COVID-19 outbreaks in care homes: early international evidence. Article in LTCcovid.org, International Long-Term Care Policy Network, CPEC-LSE, 26 June 2020. |
| Peak 7-day average of COVID-19 mortality: Worldometers.info |
| Population overall and in elderly: Seroprevalence study (based on Statistics Denmark) |

| |
|---|
| Population in nursing homes: Dataset ("Antal beboere pa plejehjem") at https://sundhedsdatastyrelsen.dk/da/tal-og-analyser/analyser-og-rapporter/sundhedsvaesenet/plejehjem |
| Proportion of elderly among nursing home residents: Statistics Denmark, dataset ("Indskrevne i pleje- og ældreboliger efter område, tid, alder og foranstaltningsart") |
| Comments:<br>Proportion of those aged ≥85 years among all elderly (≥70 years) is derived from Statistics Denmark (the whole country): 14.86% |
| **Denmark (Study: Espenhain)** |
| Seroprevalence information: Seroprevalence study |
| Age distribution of COVID-19 deaths: Riffe, T., Acosta, E., the COVerAGE-DB team, Data Resource Profile: COVerAGE-DB: a global demographic database of COVID-19 cases and deaths, International Journal of Epidemiology, Volume 50, Issue 2, April 2021, Pages 390–390f, https://doi.org/10.1093/ije/dyab027. Accessed online as 10.17605/OSF.IO/MPWJQ. |
| Total number of deaths for the primary date: Riffe, T., Acosta, E., the COVerAGE-DB team, Data Resource Profile: COVerAGE-DB: a global demographic database of COVID-19 cases and deaths, International Journal of Epidemiology, Volume 50, Issue 2, April 2021, Pages 390–390f, https://doi.org/10.1093/ije/dyab027. Accessed online as 10.17605/OSF.IO/MPWJQ. |
| COVID-19 deaths in nursing home residents: Comas-Herrera A, Zalakaín J, Litwin C, Hsu AT, Lemmon E, Henderson D and Fernández J-L (2020) Mortality associated with COVID-19 outbreaks in care homes: early international evidence. Article in LTCcovid.org, International Long-Term Care Policy Network, CPEC-LSE, 26 June 2020. |
| Peak 7-day average of COVID-19 mortality: Worldometers.info |
| Population overall and in elderly: Dataset at https://www.statistikbanken.dk/statbank5a/selectvarval/saveselections.asp |
| Population in nursing homes: Dataset ("Antal beboere pa plejehjem") at https://sundhedsdatastyrelsen.dk/da/tal-og-analyser/analyser-og-rapporter/sundhedsvaesenet/plejehjem |
| Proportion of elderly among nursing home residents: Statistics Denmark, dataset ("Indskrevne i pleje- og ældreboliger efter område, tid, alder og foranstaltningsart") |
| Comments: |
| **Dominican Republic (Study: Paulino-Ramirez)** |
| Seroprevalence information: Seroprevalence study |
| Age distribution of COVID-19 deaths: Boletín especial #65, Enfermedad por Coronavirus 2019 (COVID-19), Figura 4, situational report with data from May 22, retrieved from https://coronavirusrd.gob.do/documentos/boletines/ |
| Total number of deaths for the primary date: Boletín especial #65, Enfermedad por Coronavirus 2019 (COVID-19), situational report with data from May 22, retrieved from https://coronavirusrd.gob.do/documentos/boletines/ |
| COVID-19 deaths in nursing home residents: Unavailable, imputed |
| Peak 7-day average of COVID-19 mortality: Worldometers.info |
| Population overall and in elderly: populationpyramid.net |
| Population in nursing homes: "Living Arrangements of Older Persons: A Report on an Expanded International Dataset", United Nations (2017), Table A.III.1, retrieved from https://www.un.org/en/development/desa/population/publications/pdf/ageing/LivingArrangements.pdf |
| Proportion of elderly among nursing home residents: Imputed (average of available values) |
| **France, Ile-de-France (Study: Carrat)** |
| Seroprevalence information: Seroprevalence study |
| Age distribution of COVID-19 deaths: (1) DONNÉES DE SURVEILLANCE. SURVEILLANCE DE LA MORTALITÉ AU COURS DE L'ÉPIDÉMIE DE COVID-19 DU 2 MARS AU 31 MAI 2020 EN FRANCE, Santé publique France; (2) Point épidémio régional Ile-de-France Spécial COVID-19 28 mai 2020, Santé publique France |
| Total number of deaths for the primary date: DONNÉES DE SURVEILLANCE. SURVEILLANCE DE LA MORTALITÉ AU COURS DE L'ÉPIDÉMIE DE COVID-19 DU 2 MARS AU 31 MAI 2020 EN FRANCE, Santé publique France |
| COVID-19 deaths in nursing home residents: DONNÉES DE SURVEILLANCE. SURVEILLANCE DE LA MORTALITÉ AU COURS DE L'ÉPIDÉMIE DE COVID-19 DU 2 MARS AU 31 MAI 2020 EN FRANCE, Santé publique France |
| Peak 7-day average of COVID-19 mortality: DONNÉES DE SURVEILLANCE. SURVEILLANCE DE LA MORTALITÉ AU COURS DE L'ÉPIDÉMIE DE COVID-19 DU 2 MARS AU 31 MAI 2020 EN FRANCE, Santé publique France |
| Population overall: Webpage https://www.insee.fr/en/statistiques/3696316?p1=r11&p2=r97&lang=en&annee=2020 (accessed May 31, 2021) |
| Population in elderly: Webpage https://www.insee.fr/en/statistiques/3696316?p1=r11&p2=r97&lang=en&annee=2020 (accessed May 31, 2021) |
| Population in nursing homes: Webpage https://www.insee.fr/fr/statistiques/4124769#tableau-figure4 (accessed May 31, 2021) |
| Proportion of elderly among nursing home residents: Imputed (average of available values) |
| Comments: |

The report “Surveillance de la mortalité …” by Santé publique France, Table 5, lists the number of COVID-19 deaths in hospitalized patients and the nursing home residents’ COVID-19 deaths by May 31, per region. In Ile-de-France: 7262 hospitalized, 4368 EHPAD/EMS (total 7262+4368).
The region of Ile-de-France has a situational report from May 28, listing age distribution of deaths occurring in hospitalized patients up to May 26 in Tableau 5: 5229/6930 (age cutoff 70 years) with age information. Total number of deaths were 6958. Note that this number does not include the nursing home residents’ deaths, which is here added using the proportion of deaths in the Santé publique report, May 31.
Total number of deaths = (4368/11630)*6958+6958 = 9571. Number of deaths in elderly ≥70 years = (4368/11630)*6958*0.95+5229 = 7712.
Three regions were sampled in the study by Carrat, and two were eligible (the ones with >1000 participants 70+): Ile-de-France and Nouvelle Aquitaine. Using primary outcome (ELISA-S). Figure 2 (weighted estimates) and Suppl figure 5 (unweighted).

**France, Nouvelle Aquitaine (Study: Carrat)**

Seroprevalence information: Seroprevalence study

Age distribution of COVID-19 deaths: Point épidémio régional Nouvelle-Aquitaine Spécial COVID-19 28 mai 2020, Santé publique France

Total number of deaths for the primary date: Point épidémio régional Nouvelle-Aquitaine Spécial COVID-19 28 mai 2020, Santé publique France

COVID-19 deaths in nursing home residents: DONNÉES DE SURVEILLANCE. SURVEILLANCE DE LA MORTALITÉ AU COURS DE L'ÉPIDÉMIE DE COVID-19 DU 2 MARS AU 31 MAI 2020 EN FRANCE, Santé publique France

Peak 7-day average of COVID-19 mortality: DONNÉES DE SURVEILLANCE. SURVEILLANCE DE LA MORTALITÉ AU COURS DE L'ÉPIDÉMIE DE COVID-19 DU 2 MARS AU 31 MAI 2020 EN FRANCE, Santé publique France

Population overall: Webpage https://www.insee.fr/en/statistiques/3696316?p1=r11&p2=r97&lang=en&annee=2020 (accessed May 31, 2021)

Population in elderly: Webpage https://www.insee.fr/en/statistiques/3696316?p1=r11&p2=r97&lang=en&annee=2020 (accessed May 31, 2021)

Population in nursing homes: Webpage https://www.insee.fr/fr/statistiques/4294178#tableau-figure2 (accessed May 31, 2021)

Proportion of elderly among nursing home residents: Imputed (average of available values)

Comments:
The report “Surveillance de la mortalité …” by Santé publique France, Table 5, lists the number of COVID-19 deaths in hospitalized patients and the nursing home residents’ COVID-19 deaths, per region. Nouvelle Aquitaine: 406 hospitalized, 129 EHPAD/EMS (total 406+129).
The region of Nouvelle Aquitaine has a situational report from May 28, listing age distribution of all deaths up to May 25 in Tableau 7: 409/448 (age cutoff 65 years).
Population in nursing homes: Derived from the Insee webpage Figure 2, Number of dependent seniors (2016), n=331,200. In text: "in 2016, 8 out of 10 dependent elderly people lived at home" [and the others in institutions].
Three regions were sampled in the study by Carrat, and two were eligible (the ones with >1000 participants 70+): Ile-de-France and Nouvelle Aquitaine. Using primary outcome (ELISA-S). Figure 2 (weighted estimates) and Suppl figure 5 (unweighted).

**France (Study: Warszawski / INSERM)**

Seroprevalence information: Seroprevalence study

Age distribution of COVID-19 deaths: Point épidémiologique hebdomadaire du 03 décembre 2020, Santé publique France, Tableau 2 and Tableau 4, retrieved from https://www.santepubliquefrance.fr/recherche/#search=COVID%2019%20%20%20point%20epidemiologique&publications=donn%C3%A9es&regions=National&sort=date

Total number of deaths for the primary date: Point épidémiologique hebdomadaire du 03 décembre 2020, Santé publique France, Tableau 2 and Tableau 4, retrieved from https://www.santepubliquefrance.fr/recherche/#search=COVID%2019%20%20%20point%20epidemiologique&publications=donn%C3%A9es&regions=National&sort=date

COVID-19 deaths in nursing home residents: Point épidémiologique hebdomadaire du 03 décembre 2020, Santé publique France, Tableau 2 and Tableau 4, retrieved from https://www.santepubliquefrance.fr/recherche/#search=COVID%2019%20%20%20point%20epidemiologique&publications=donn%C3%A9es&regions=National&sort=date

Peak 7-day average of COVID-19 mortality: Worldometers.info

Population overall and in elderly: INSEE (Age structure of the population on 1st January 2020, metropolitan France), dataset at https://www.insee.fr/en/statistiques/5015921?sommaire=5015923

Population in nursing homes: INSEE (Structures d'hébergement pour personnes âgées, December 31, 2016), dataset at https://www.insee.fr/fr/statistiques/3676717?sommaire=3696937

Proportion of elderly among nursing home residents: Imputed (average of available values)

| |
|---|
| Comments:<br>The number of deaths in elderly is derived from Tableau 4 (deaths occurring in hospital) and Tableau 2 (deaths in care homes) assuming that 98% of the 16,664 deaths in care homes were among persons 65 years and older according to our prespecification. |
| **Hungary (Study: Merkely)** |
| Seroprevalence information: Personal communication Z. Vokó |
| Age distribution of COVID-19 deaths: Early phase of the COVID-19 outbreak in Hungary and post-lockdown scenarios<br>Gergely Röst, Ferenc A. Bartha, Norbert Bogya, Péter Boldog, Attila Dénes, Tamás Ferenci, Krisztina J. Horváth, Attila Juhász, Csilla Nagy, Tamás Tekeli, Zsolt Vizi, Beatrix Oroszi. (Version 1) medRxiv 2020.06.02.20119313; doi: https://doi.org/10.1101/2020.06.02.20119313 |
| Total number of deaths for the primary date: Worldometers.info (accessed June 1, 2021) |
| COVID-19 deaths in nursing home residents: Comas-Herrera A, Zalakaín J, Litwin C, Hsu AT, Lane N and Fernández J-L (2020) Mortality associated with COVID-19 outbreaks in care homes: early international evidence. Article in LTCcovid.org, International Long-Term Care Policy Network, CPEC-LSE, 21 May 2020. |
| Peak 7-day average of COVID-19 mortality: Worldometers.info |
| Population overall and in elderly: populationpyramid.net |
| Population in nursing homes: Comas-Herrera A, Zalakaín J, Lemmon E, Henderson D, Litwin C, Hsu AT, Schmidt AE, Arling G and Fernández J-L (2020) Mortality associated with COVID-19 in care homes: international evidence. Article in LTCcovid.org, International Long-Term Care Policy Network, CPEC-LSE, 14 October. (page 10, in text) |
| Proportion of elderly among nursing home residents: Comas-Herrera A, Zalakaín J, Lemmon E, Henderson D, Litwin C, Hsu AT, Schmidt AE, Arling G and Fernández J-L (2020) Mortality associated with COVID-19 in care homes: international evidence. Article in LTCcovid.org, International Long-Term Care Policy Network, CPEC-LSE, 14 October. (page 10, in text) |
| **Iceland (Study: Gudbjartsson)** |
| Seroprevalence information: Seroprevalence study and https://www.ruv.is/frett/2020/04/20/annad-andlat-ur-covid-19-a-hjukrunarheimilinu-bergi (5 infected nursing home residents) |
| Age distribution of COVID-19 deaths: Webpage, https://www.covid.is/data-old (last updated: June 14, 2020; accessed: May 29, 2021) |
| Total number of deaths for the primary date: Webpage, https://www.covid.is/data-old (last updated: June 14, 2020; accessed: May 29, 2021) |
| COVID-19 deaths in nursing home residents: Wikipedia, https://en.wikipedia.org/wiki/COVID-19_pandemic_in_Iceland (accessed: May 29, 2021) |
| Peak 7-day average of COVID-19 mortality: Worldometers.info |
| Population overall and in elderly: populationpyramid.net |
| Population in nursing homes: Dataset "Occupants of retirement homes and nursing homes and wards by type of institution 1993-2010" (selection: 2010, In nursing homes) retrieved from https://px.hagstofa.is/pxen/pxweb/en/Samfelag/Samfelag__felagsmal__aldradir/HEI03004.px |
| Proportion of elderly among nursing home residents: Imputed (average of available values) |
| Comments:<br>The seroprevalence study excluded previously COVID-19-diagnosed persons, and presents prevalence based on antibody tests and PCR tests. The estimate given is the number of infected Icelanders of age 70+ (i.e., the absolute number, not seroprevalence), Table S7 (n=165.4). In the calculations to follow, we subtract 5 infections in nursing homes, see website source. Gudbjartsson's estimated IFR is 4.4% (1.9%, 8.3%). Since no confidence intervals are given for the number of infected persons (or seroprevalence), their confidence intervals for IFR are used in Figure 1.<br>Mortality peak date is the midpoint between March 27 and April 23. |
| **India (Study: Murhekar)** |
| Seroprevalence information: Seroprevalence study |
| Age distribution of COVID-19 deaths: Press conference by India's health secretary October 13, referenced at https://science.thewire.in/health/india-covid-19-mortality-comorbidities-age-health-ministry/ |
| Total number of deaths for the primary date: Worldometers.info (accessed June 1, 2021) |
| COVID-19 deaths in nursing home residents: Unavailable, imputed |
| Peak 7-day average of COVID-19 mortality: Worldometers.info |
| Population overall and in elderly: populationpyramid.net |
| Population in nursing homes: "Report on old age facilities in India", Tata Trusts, Samarth, and United Nations Population Fund (2018), retrieved from https://www.tatatrusts.org/upload/pdf/report-on-old-age-facilities-in-india.pdf |
| **India [Tamil Nadu] (Study: Malani)** |

| |
|---|
| Seroprevalence information: Seroprevalence study |
| Age distribution of COVID-19 deaths: Seroprevalence study |
| Total number of deaths for the primary date: Seroprevalence study |
| COVID-19 deaths in nursing home residents: Unavailable, imputed |
| Peak 7-day average of COVID-19 mortality: Wikipedia |
| Population overall and in elderly: Seroprevalence study (Census 2011) |
| Population in nursing homes: “Report on old age facilities in India”, Tata Trusts, Samarth, and United Nations Population Fund (2018), retrieved from https://www.tatatrusts.org/upload/pdf/report-on-old-age-facilities-in-india.pdf (proportion in India applied to population in Tamil Nadu) |
| Proportion of elderly among nursing home residents: Imputed (average of available values) |
| Comments:<br>Mortality peak is the midpoint of Sept 15, 2020, and Sept 19, 2020. |
| **Italy (Study: Istat)** |
| Seroprevalence information: Seroprevalence study |
| Age distribution of COVID-19 deaths: Epidemia COVID-19, Aggiornamento nazionale, 23 giugno 2020 – ore 11:00, DATA PUBBLICAZIONE: 26 GIUGNO 2020, ISS, retrieved from https://www.epicentro.iss.it/coronavirus/bollettino/Bollettino-sorveglianza-integrata-COVID-19_23-giugno-2020.pdf |
| Total number of deaths for the primary date: Worldometers.info |
| COVID-19 deaths in nursing home residents: (1) Comas-Herrera A, Zalakaín J, Litwin C, Hsu AT, Lemmon E, Henderson D and Fernández J-L (2020) Mortality associated with COVID-19 outbreaks in care homes: early international evidence. Article in LTCcovid.org, International Long-Term Care Policy Network, CPEC-LSE, 26 June 2020 [for the absolute number of nursing home residents estimated to have died by May 5, 2020, n=9212]; (2) Worldometers.info [for the total number of deaths at the corresponding date, n=29389] |
| Peak 7-day average of COVID-19 mortality: Worldometers.info |
| Population overall and in elderly: populationpyramid.net |
| Population in nursing homes: Comas-Herrera A, Zalakaín J, Litwin C, Hsu AT, Lemmon E, Henderson D and Fernández J-L (2020) Mortality associated with COVID-19 outbreaks in care homes: early international evidence. Article in LTCcovid.org, International Long-Term Care Policy Network, CPEC-LSE, 26 June 2020 |
| Proportion of elderly among nursing home residents: Imputed (average of available values) |
| Comments:<br>COVID-19 deaths in nursing home residents, calculated from LTC Policy Network report June 26, Table 2: 3.1% of 297,158 residents were estimated to have died of COVID-19 at that point. |
| **Netherlands (Study: Vos) – This study is not included in the main analysis, but in one sensitivity analysis** |
| Seroprevalence information: Seroprevalence study |
| Age distribution of COVID-19 deaths: Epidemiologische situatie COVID-19 in Nederland. Rijksinstituut voor Volksgezondheid en Milieu – RIVM. 21 juni 2020, 10:00. Retrieved from: https://www.rivm.nl/coronavirus-covid-19/actueel/wekelijkse-update-epidemiologische-situatie-covid-19-in-nederland |
| Total number of deaths for the primary date: Epidemiologische situatie COVID-19 in Nederland. Rijksinstituut voor Volksgezondheid en Milieu – RIVM. 21 juni 2020, 10:00. Retrieved from: https://www.rivm.nl/coronavirus-covid-19/actueel/wekelijkse-update-epidemiologische-situatie-covid-19-in-nederland |
| COVID-19 deaths in nursing home residents: Rijksinstituut voor Volksgezondheid en Milieu (RIVM) “open data set on nursing home care”, retrieved 2021-12-17 from https://coronadashboard.government.nl/verantwoording#nursing-homes |
| Peak 7-day average of COVID-19 mortality: Worldometers.info |
| Population overall and in elderly: Statistics Netherlands (CBS), Population pyramid: Age composition in the Nederlands 2021. Retrieved from: https://www.cbs.nl/en-gb/visualisations/dashboard-population/population-pyramid |
| Population in nursing homes: Statistics Netherlands (CBS), Households; size, composition, position in the household, 1 January (2021). Retrieved from: https://www.cbs.nl/en-gb/society/population |
| Proportion of elderly among nursing home residents: Imputed (average of available values) |
| Comments: |
| **Qatar (Study: Abu-Raddad)** |
| Seroprevalence information: Seroprevalence study |
| Age distribution of COVID-19 deaths: Seroprevalence study |
| Total number of deaths for the primary date: Worldometers.info (accessed June 1, 2021) |
| COVID-19 deaths in nursing home residents: Unavailable, imputed |
| Peak 7-day average of COVID-19 mortality: Worldometers.info |

| |
|---|
| Population overall and in elderly: populationpyramid.net |
| Population in nursing homes: ”Living Arrangements of Older Persons: A Report on an Expanded International Dataset”, United Nations (2017), Table A.III.1, retrieved from https://www.un.org/en/development/desa/population/publications/pdf/ageing/LivingArrangements.pdf |
| Proportion of elderly among nursing home residents: Imputed (average of available values) |
| Comments:<br>Confidence intervals for adjusted seroprevalence not reported in study, and so calculated using the inferred adjusted number of seropositive individuals.<br>Population in nursing homes: The number for females (0.5% of elderly living in institutions) selected since the definition of institutional living included other collective housing than elderly homes, and the male population includes a large number of workers living in collective quarters.<br>Mortality peak date is the midpoint between June 21, 2020, and July 10, 2020. |
| **Spain (Study: Ministerio de Sanidad and Instituto de Salud Carlos III)** |
| Seroprevalence information: Seroprevalence study |
| Age distribution of COVID-19 deaths: Dataset "casos_hosp_uci_def_sexo_edad_provres.csv" retrieved from https://cnecovid.isciii.es/covid19/#documentación-y-datos, accessed 2021-06-23 |
| Total number of deaths for the primary date: Dataset "casos_hosp_uci_def_sexo_edad_provres.csv" retrieved from https://cnecovid.isciii.es/covid19/#documentación-y-datos, accessed 2021-06-23 |
| COVID-19 deaths in nursing home residents: Comas-Herrera A, Zalakaín J, Lemmon E, Henderson D, Litwin C, Hsu AT, Schmidt AE, Arling G, Kruse F and Fernández J-L (2020) Mortality associated with COVID-19 in care homes: international evidence. Article in LTCcovid.org, International Long-Term Care Policy Network, CPEC-LSE, 1st February 2021. |
| Peak 7-day average of COVID-19 mortality: Worldometers.info |
| Population overall and in elderly: populationpyramid.net |
| Population in nursing homes: Webpage, http://envejecimientoenred.es/una-nueva-estimacion-de-poblacion-en-residencias-de-mayores/ (July 20, 2020; accessed May 29, 2021) |
| Proportion of elderly among nursing home residents: Webpage, http://envejecimientoenred.es/una-nueva-estimacion-de-poblacion-en-residencias-de-mayores/ (July 20, 2020; accessed May 29, 2021) |
| **United Kingdom (Study: UK Biobank)** |
| Seroprevalence information: Seroprevalence study |
| Age distribution of COVID-19 deaths: Deaths registered weekly in England and Wales, provisional, Office of National Statistics, dataset (sheet "UK - Covid-19 - Weekly reg") retrieved from https://www.ons.gov.uk/peoplepopulationandcommunity/birthsdeathsandmarriages/deaths/datasets/weeklyprovisionalfiguresondeathsregisteredinenglandandwales (accessed February 19, 2021). Note that this dataset includes numbers for the United Kingdom, not only England and Wales. |
| Total number of deaths for the primary date: Deaths registered weekly in England and Wales, provisional, Office of National Statistics, dataset (sheet "UK - Covid-19 - Weekly reg") retrieved from https://www.ons.gov.uk/peoplepopulationandcommunity/birthsdeathsandmarriages/deaths/datasets/weeklyprovisionalfiguresondeathsregisteredinenglandandwales (accessed February 19, 2021). Note that this dataset includes numbers for the United Kingdom, not only England and Wales. |
| COVID-19 deaths in nursing home residents: Comas-Herrera A, Zalakaín J, Lemmon E, Henderson D, Litwin C, Hsu AT, Schmidt AE, Arling G and Fernández J-L (2020) Mortality associated with COVID-19 in care homes: international evidence. Article in LTCcovid.org, International Long-Term Care Policy Network, CPEC-LSE, 14 October |
| Peak 7-day average of COVID-19 mortality: Worldometers.info |
| Population overall and in elderly: Population estimates for the UK, England and Wales, Scotland and Northern Ireland: mid-2019, using April 2020 local authority district codes, Office for National Statistics (sheet “MYE1”), retrieved from https://www.ons.gov.uk/peoplepopulationandcommunity/populationandmigration/populationestimates/datasets/populationestimatesforukenglandandwalesscotlandandnorthernireland, "Mid-2019: April 2020 local authority district codes" |
| Population in nursing homes: Comas-Herrera A, Zalakaín J, Lemmon E, Henderson D, Litwin C, Hsu AT, Schmidt AE, Arling G and Fernández J-L (2020) Mortality associated with COVID-19 in care homes: international evidence. Article in LTCcovid.org, International Long-Term Care Policy Network, CPEC-LSE, 14 October (Table 2, column “% of pop living in care homes”) |
| Proportion of elderly among nursing home residents: Bowman et al 2004, Age and Ageing 2004; 33: 561–566, doi:10.1093/ageing/afh177 |
| Comments:<br>The alternative sampling period May 27, 2020, to July 6, 2020, is discarded according to eligibility criteria (lower seroprevalence). |
| **United Kingdom [England] (Study: Ward)** |

| |
|---|
| Seroprevalence information: Seroprevalence study |
| Age distribution of COVID-19 deaths: Deaths registered weekly in England and Wales, provisional, Office of National Statistics, dataset (sheet "UK - Covid-19 - Weekly occurrences") retrieved from https://www.ons.gov.uk/peoplepopulationandcommunity/birthsdeathsandmarriages/deaths/datasets/weeklyprovisionalfiguresondeathsregisteredinenglandandwales (accessed February 19, 2021). The age distribution for England and Wales is used. |
| Total number of deaths for the primary date: Deaths registered weekly in England and Wales, provisional, Office of National Statistics, dataset (sheet "UK - Covid-19 - Weekly occurrences") retrieved from https://www.ons.gov.uk/peoplepopulationandcommunity/birthsdeathsandmarriages/deaths/datasets/weeklyprovisionalfiguresondeathsregisteredinenglandandwales (accessed February 19, 2021). |
| COVID-19 deaths in nursing home residents: Dataset ("julydeathsinvolvingcovid19inthecaresectordataset02072020155122.xlsx", sheet: “Table 1”), retrieved from https://www.ons.gov.uk/peoplepopulationandcommunity/birthsdeathsandmarriages/deaths/datasets/deathsinvolvingcovid19inthecaresectorenglandandwales (accessed February 19, 2021) |
| Peak 7-day average of COVID-19 mortality: Deaths registered weekly in England and Wales, provisional, Office of National Statistics, dataset (sheet "UK - Covid-19 - Daily occurrences") retrieved from https://www.ons.gov.uk/peoplepopulationandcommunity/birthsdeathsandmarriages/deaths/datasets/weeklyprovisionalfiguresondeathsregisteredinenglandandwales (accessed February 19, 2021). |
| Population overall and in elderly: Population estimates for the UK, England and Wales, Scotland and Northern Ireland: mid-2019, using April 2020 local authority district codes, Office for National Statistics (sheet “MYE1”), retrieved from https://www.ons.gov.uk/peoplepopulationandcommunity/populationandmigration/populationestimates/datasets/populationestimatesforukenglandandwalesscotlandandnorthernireland, "Mid-2019: April 2020 local authority district codes" |
| Population in nursing homes: Comas-Herrera A, Zalakaín J, Lemmon E, Henderson D, Litwin C, Hsu AT, Schmidt AE, Arling G and Fernández J-L (2020) Mortality associated with COVID-19 in care homes: international evidence. Article in LTCcovid.org, International Long-Term Care Policy Network, CPEC-LSE, 14 October (Table 2, column “% of pop living in care homes”) |
| Proportion of elderly among nursing home residents: Dataset at http://www.nomisweb.co.uk/census/2011/all_tables?release=3.4, "DC4210EWIa", England |
| Comments: The age distribution of COVID-19 mortality for England and Wales is used. |
| **United Kingdom [England and Wales] (Study: Public Health England)** |
| Seroprevalence information: Seroprevalence study |
| Age distribution of COVID-19 deaths: Deaths registered weekly in England and Wales, provisional, Office of National Statistics, dataset (sheet "UK - Covid-19 - Weekly occurrences") retrieved from https://www.ons.gov.uk/peoplepopulationandcommunity/birthsdeathsandmarriages/deaths/datasets/weeklyprovisionalfiguresondeathsregisteredinenglandandwales (accessed February 19, 2021). |
| Total number of deaths for the primary date: Deaths registered weekly in England and Wales, provisional, Office of National Statistics, dataset (sheet "UK - Covid-19 - Weekly occurrences") retrieved from https://www.ons.gov.uk/peoplepopulationandcommunity/birthsdeathsandmarriages/deaths/datasets/weeklyprovisionalfiguresondeathsregisteredinenglandandwales (accessed February 19, 2021). |
| COVID-19 deaths in nursing home residents: Dataset ("julydeathsinvolvingcovid19inthecaresectordataset02072020155122.xlsx", sheet: “Table 1”), retrieved from https://www.ons.gov.uk/peoplepopulationandcommunity/birthsdeathsandmarriages/deaths/datasets/deathsinvolvingcovid19inthecaresectorenglandandwales (accessed February 19, 2021) |
| Peak 7-day average of COVID-19 mortality: Deaths registered weekly in England and Wales, provisional, Office of National Statistics, dataset (sheet "UK - Covid-19 - Daily occurrences") retrieved from https://www.ons.gov.uk/peoplepopulationandcommunity/birthsdeathsandmarriages/deaths/datasets/weeklyprovisionalfiguresondeathsregisteredinenglandandwales (accessed February 19, 2021). |
| Population overall and in elderly: Population estimates for the UK, England and Wales, Scotland and Northern Ireland: mid-2019, using April 2020 local authority district codes, Office for National Statistics (sheet “MYE1”), retrieved from https://www.ons.gov.uk/peoplepopulationandcommunity/populationandmigration/populationestimates/datasets/populationestimatesforukenglandandwalesscotlandandnorthernireland, "Mid-2019: April 2020 local authority district codes" |
| Population in nursing homes: Comas-Herrera A, Zalakaín J, Lemmon E, Henderson D, Litwin C, Hsu AT, Schmidt AE, Arling G and Fernández J-L (2020) Mortality associated with COVID-19 in care homes: international evidence. Article in LTCcovid.org, International Long-Term Care Policy Network, CPEC-LSE, 14 October (Table 2, column “% of pop living in care homes”, England) |
| Proportion of elderly among nursing home residents: Dataset at http://www.nomisweb.co.uk/census/2011/all_tables?release=3.4, "DC4210EWIa", England and Wales |
| Comments:<br>First of two sampling periods, chosen according to eligibility criteria (highest seroprevalence). |

| |
|---|
| **United Kingdom [Greater Glasgow and Clyde, Scotland] (Study: Hughes)** |
| Seroprevalence information: Seroprevalence study |
| Age distribution of COVID-19 deaths: Weekly deaths by health board, age group, sex and cause, 2020 and 2021 , dataset retrieved from https://www.nrscotland.gov.uk/files/statistics/covid19/weekly-deaths-by-sex-age-group-health-board-2020-2021.xlsx (accessed February 19, 2021; date range specified according to primary date) |
| Total number of deaths for the primary date: Weekly deaths by health board, age group, sex and cause, 2020 and 2021, dataset retrieved from https://www.nrscotland.gov.uk/files/statistics/covid19/weekly-deaths-by-sex-age-group-health-board-2020-2021.xlsx (accessed February 19, 2021; date range specified according to primary date) |
| COVID-19 deaths in nursing home residents: Weekly deaths by area and location, 2020 and 2021 (sheet: “HB COVID”), dataset retrieved from https://www.nrscotland.gov.uk/files/statistics/covid19/weekly-deaths-by-location-health-board-council-area-2020-2021.xlsx |
| Peak 7-day average of COVID-19 mortality: Deaths involving coronavirus (COVID-19) in Scotland (Table 1 (2020), dataset retrieved from https://www.nrscotland.gov.uk/statistics-and-data/statistics/statistics-by-theme/vital-events/general-publications/weekly-and-monthly-data-on-births-and-deaths/deaths-involving-coronavirus-covid-19-in-scotland |
| Population overall and in elderly: "Mid-2019 population estimates Scotland" (sheet: “Table 3”), dataset retrieved from https://www.nrscotland.gov.uk/statistics-and-data/statistics/statistics-by-theme/population/population-estimates/mid-year-population-estimates/mid-2019 |
| Population in nursing homes: Scottish Care Home Census, Public Health Scotland (PHS), "Care Home Census Data Tables", (sheet: “Table 3”), dataset retrieved from https://beta.isdscotland.org/find-publications-and-data/health-and-social-care/social-and-community-care/care-home-census-for-adults-in-scotland/ |
| Proportion of elderly among nursing home residents: Scottish Care Home Census, Public Health Scotland (PHS), "Care Home Census Data Tables", (sheet: “Table 8”), dataset retrieved from https://beta.isdscotland.org/find-publications-and-data/health-and-social-care/social-and-community-care/care-home-census-for-adults-in-scotland/ |
| Comments:<br>NHSGGC (base for seroprevalence study sampling) covers the following local authorities: Inverclyde, Renfrewshire, East Renfrewshire, Glasgow City, East Dunbartonshire and West Dunbartonshire.<br>COVID-19 death statistics for nursing home residents do not include deaths occurring in hospital, and so was corrected with a factor of 1.225 (the median of the ratio of deaths in nursing home residents / deaths occurring in nursing homes, in the European countries with such data in the Long-term Care Policy Network report October 14, Comas-Herrera et al). |
| **United States of America (Study: Anand)** |
| Seroprevalence information: Seroprevalence study |
| Age distribution of COVID-19 deaths: Dataset retrieved from https://data.cdc.gov/NCHS/Provisional-COVID-19-Death-Counts-by-Sex-Age-and-W/vsak-wrfu (accessed February 19, 2021) |
| Total number of deaths for the primary date: Worldometers.info (accessed June 1, 2021) |
| COVID-19 deaths in nursing home residents: Webpage, https://www.kff.org/policy-watch/this-week-in-coronavirus-july-10-to-july-16/ (accessed May 5, 2021) |
| Peak 7-day average of COVID-19 mortality: Worldometers.info |
| Population overall and in elderly: "Annual Estimates of the Resident Population for Selected Age Groups by Sex: April 1, 2010 to July 1, 2019", dataset retrieved from https://www.census.gov/data/tables/time-series/demo/popest/2010s-national-detail.html |
| Population in nursing homes: Comas-Herrera A, Zalakaín J, Lemmon E, Henderson D, Litwin C, Hsu AT, Schmidt AE, Arling G and Fernández J-L (2020) Mortality associated with COVID-19 in care homes: international evidence. Article in LTCcovid.org, International Long-Term Care Policy Network, CPEC-LSE, 14 October (Table 2, column “% of pop living in care homes”) |
| Proportion of elderly among nursing home residents: Nursing Home Data Compendium 2015 Edition, Department of Health & Human Services USA (Table 3.11.d), retrieved from https://www.cms.gov/Medicare/Provider-Enrollment-and-Certification/CertificationandComplianc/Downloads/nursinghomedatacompendium_508-2015.pdf |
| Comments: Deaths in long-term care facilities based on data from 42 states. Total is derived from extracted percentage and long-term care deaths. |
| **United States of America (Study: Kalish)** |
| Seroprevalence information: Seroprevalence study and webpages https://covid.cdc.gov/covid-data-tracker/#demographicsovertime and https://www.kff.org/policy-watch/this-week-in-coronavirus-june-11-to-june-17/ (accessed May 5, 2021; used to calculate the number of cases in community-dwelling elderly, see Comments) |
| Age distribution of COVID-19 deaths: Dataset retrieved from https://data.cdc.gov/NCHS/Provisional-COVID-19-Death-Counts-by-Sex-Age-and-W/vsak-wrfu (accessed February 19, 2021) |
| Total number of deaths for the primary date: Worldometers.info (accessed June 1, 2021) |
| COVID-19 deaths in nursing home residents: Webpage, https://www.kff.org/policy-watch/this-week-in-coronavirus-june-18-to-june-25/ (Accessed May 5, 2021) |

| |
|---|
| Peak 7-day average of COVID-19 mortality: Worldometers.info |
| Population overall and in elderly: "Annual Estimates of the Resident Population for Selected Age Groups by Sex: April 1, 2010 to July 1, 2019", dataset retrieved from https://www.census.gov/data/tables/time-series/demo/popest/2010s-national-detail.html |
| Population in nursing homes: Comas-Herrera A, Zalakaín J, Lemmon E, Henderson D, Litwin C, Hsu AT, Schmidt AE, Arling G and Fernández J-L (2020) Mortality associated with COVID-19 in care homes: international evidence. Article in LTCcovid.org, International Long-Term Care Policy Network, CPEC-LSE, 14 October (Table 2, column "% of pop living in care homes") |
| Proportion of elderly among nursing home residents: Nursing Home Data Compendium 2015 Edition, Department of Health & Human Services USA (Table 3.11.d), retrieved from https://www.cms.gov/Medicare/Provider-Enrollment-and-Certification/CertificationandComplianc/Downloads/nursinghomedatacompendium_508-2015.pdf |
| Comments:<br>Deaths in long-term care facilities based on data from 42 states. Total is derived from extracted percentage and long-term care deaths.<br>The study specifically targeted people not previously diagnosed with COVID-19, and so the total number of infected was corrected. We calculated the number of cases in community-dwelling elderly up to June 20 by retrieving the cumulative number of cases in elderly (≥65) from the COVID-19 Data Tracker at the US CDC (455,585) and subtracting the cumulative number of cases in nursing homes from KFF (240,138), resulting in 215,447 cases. This number was added to the number of infected community-dwelling elderly. |

**Appendix Table 3a.** Reports not included, which sampled or potentially sampled ≥1000 participants ≥70 years of age and were conducted in 2020.

| DOI or URL | Country | Reason for exclusion |
|---|---|---|
| 10.1101/2020.08.10.20171942v1 | Brazil | COVID-19 mortality data unavailable for elderly at study location |
| https://pubmed.ncbi.nlm.nih.gov/33883260/ | China | COVID-19 mortality data unavailable for elderly at study location |
| 10.1016/j.ijid.2021.05.043 | India | COVID-19 mortality data unavailable for elderly at study location |
| 10.1007/s10654-021-00749-1 | Israel | COVID-19 mortality data unavailable for elderly at study location |
| 10.3390/diagnostics11030483 | Italy | COVID-19 mortality data unavailable for elderly at study location |
| 10.2144/fsoa-2020-0203 | Italy | COVID-19 mortality data unavailable for elderly at study location |
| 10.19191/EP20.5-6.S2.119 | Italy | COVID-19 mortality data unavailable for elderly at study location |
| 10.31662/jmaj.2020-0094 | Japan | COVID-19 mortality data unavailable for elderly at study location |
| 10.3390/v13081648 | Russia | COVID-19 mortality data unavailable for elderly at study location |
| https://www.rtve.es/noticias/20200617/test-serologicos-torrejon-ardoz-arrojan-2018-habitantes-tiene-anticuerpos-coronavirus/2018787.shtml | Spain | COVID-19 mortality data unavailable for elderly at study location |
| 10.1016/j.xkme.2021.01.002 | USA | COVID-19 mortality data unavailable for elderly at study location |
| 10.1016/j.diagmicrobio.2020.115128 | USA | COVID-19 mortality data unavailable for elderly at study location |
| 10.1111/1753-6405.13155 | Australia | No seroprevalence estimate for elderly |
| 10.1038/s41591-020-0949-6 | China | No seroprevalence estimate for elderly |
| https://www.ifo.de/en/publikationen/2020/monograph-authorship/die-deutschen-und-corona | Germany | No seroprevalence estimate for elderly |
| 10.1093/trstmh/trab109 | India | No seroprevalence estimate for elderly |
| https://www.researchsquare.com/article/rs-80259/v1 | India | No seroprevalence estimate for elderly |
| 10.2188/jea.JE20210324 | Japan | No seroprevalence estimate for elderly (only for a time period in 2021, ineligible) |
| 10.1038/s41598-021-89236-x | Spain | No seroprevalence estimate for elderly |
| https://www.sll.se/verksamhet/halsa-och-vard/nyheter-halsa-och-vard/2020/09/25-september-lagesrapport-om-arbetet-med-det-nya-coronaviruset/ | Sweden | No seroprevalence estimate for elderly; not general population |
| https://www.sll.se/verksamhet/halsa-och-vard/nyheter-halsa-och-vard/2020/09/29-september-lagesrapport-om-covid-19/ | Sweden | No seroprevalence estimate for elderly; not general population |
| https://www.sll.se/verksamhet/halsa-och-vard/nyheter-halsa-och-vard/2020/10/6-oktober-lagesrapport-om-covid-19/ | Sweden | No seroprevalence estimate for elderly; not general population |
| https://www.sll.se/verksamhet/halsa-och-vard/nyheter-halsa-och- | Sweden | No seroprevalence estimate for elderly; not general population |

| | | |
|---|---|---|
| vard/2020/10/13-oktober-lagesrapport-om-covid-19/ | | |
| https://www.sll.se/verksamhet/halsa-och-vard/nyheter-halsa-och-vard/2020/10/20-oktober-lagesrapport-om-covid-19/ | Sweden | No seroprevalence estimate for elderly; not general population |
| https://www.sll.se/verksamhet/halsa-och-vard/nyheter-halsa-och-vard/2020/10/27-oktober-lagesrapport-om-covid-19/ | Sweden | No seroprevalence estimate for elderly; not general population |
| https://www.sll.se/verksamhet/halsa-och-vard/nyheter-halsa-och-vard/2020/11/3-november-lagesrapport-om-covid-19/ | Sweden | No seroprevalence estimate for elderly; not general population |
| https://www.sll.se/verksamhet/halsa-och-vard/nyheter-halsa-och-vard/2020/11/10-november-lagesrapport-om-covid-19/ | Sweden | No seroprevalence estimate for elderly; not general population |
| https://www.sll.se/verksamhet/halsa-och-vard/nyheter-halsa-och-vard/2020/11/17-november-lagesrapport-om-covid-19/ | Sweden | No seroprevalence estimate for elderly; not general population |
| https://www.sll.se/verksamhet/halsa-och-vard/nyheter-halsa-och-vard/2020/11/24-november-lagesrapport-om-covid-19/ | Sweden | No seroprevalence estimate for elderly; not general population |
| https://www.sll.se/verksamhet/halsa-och-vard/nyheter-halsa-och-vard/2020/12/1-december-lagesrapport-om-covid-19/ | Sweden | No seroprevalence estimate for elderly; not general population |
| https://www.sll.se/verksamhet/halsa-och-vard/nyheter-halsa-och-vard/2020/12/8-december-lagesrapport-om-covid-19/ | Sweden | No seroprevalence estimate for elderly; not general population |
| https://www.sll.se/verksamhet/halsa-och-vard/nyheter-halsa-och-vard/2020/12/15-december-lagesrapport-om-covid-19/ | Sweden | No seroprevalence estimate for elderly; not general population |
| https://www.sll.se/verksamhet/halsa-och-vard/nyheter-halsa-och-vard/2020/12/22-december-lagesrapport-om-covid-19/ | Sweden | No seroprevalence estimate for elderly; not general population |
| https://www.sll.se/verksamhet/halsa-och-vard/nyheter-halsa-och-vard/2020/12/29-december-lagesrapport-om-covid-19/ | Sweden | No seroprevalence estimate for elderly; not general population |
| https://www.ons.gov.uk/peoplepopulationandcommunity/healthandsocialcare/conditionsanddiseases/articles/coronaviruscovid19infectionsinthecommunityinengland/december2020 | UK | No seroprevalence estimate for elderly (only for a time period in 2021, ineligible) |
| 10.1016/j.tmrv.2021.07.001 | USA | No seroprevalence estimate for elderly |
| 10.1093/infdis/jiab514 | USA | No seroprevalence estimate for elderly |
| 10.3390/jcm10194341 | USA | No seroprevalence estimate for elderly; not explicitly aiming to generate sample reflecting general population |
| 10.1016/j.annepidem.2020.06.004 | USA | No seroprevalence estimate for elderly |
| 10.1038/s41586-020-2912-6 | USA | No seroprevalence estimate for elderly |

| | | |
|---|---|---|
| 10.1001/jama.2020.18598 | USA | No seroprevalence estimate for elderly |
| https://ejmcm.com/article_9453.html | Uzbekistan | No seroprevalence estimate for elderly; not general population |
| 10.1016/j.cmi.2020.09.044 | China | Targets high-risk participants / Does not explicitly aim to generate sample reflecting general population |
| 10.1111/all.14622 | China | Targets high-risk participants / Does not explicitly aim to generate sample reflecting general population |
| 10.1101/2020.08.10.20171850v1 | Denmark | Targets high-risk participants / Does not explicitly aim to generate sample reflecting general population |
| 10.1016/j.revmed.2021.10.330 | France | Targets high-risk participants / Does not explicitly aim to generate sample reflecting general population |
| 10.1101/2021.03.19.21253429 | India | Targets high-risk participants / Does not explicitly aim to generate sample reflecting general population |
| 10.1001/jamanetworkopen.2021.15699 | Italy | Targets high-risk participants / Does not explicitly aim to generate sample reflecting general population |
| 10.1101/2021.01.05.21249247 | Qatar | Targets high-risk participants / Does not explicitly aim to generate sample reflecting general population |
| 10.1093/ageing/afab096 | Spain | Targets high-risk participants / Does not explicitly aim to generate sample reflecting general population |
| 10.1016/S0140-6736(21)00675-9 | UK | Targets high-risk participants / Does not explicitly aim to generate sample reflecting general population |
| 10.1371/journal.pone.0252818 | USA | Targets high-risk participants / Does not explicitly aim to generate sample reflecting general population |
| https://academic.oup.com/jid/advance-article/doi/10.1093/infdis/jiab200/6219118 | USA | Targets high-risk participants / Does not explicitly aim to generate sample reflecting general population |
| 10.1093/cid/ciab519 | USA | Targets high-risk participants / Does not explicitly aim to generate sample reflecting general population |
| 10.1001/jamainternmed.2021.0366 | USA | Targets high-risk participants / Does not explicitly aim to generate sample reflecting general population |
| 10.1016/j.mayocp.2021.03.015 | USA | Targets high-risk participants / Does not explicitly aim to generate sample reflecting general population |
| 10.1093/cid/ciaa1684 | USA | Targets high-risk participants / Does not explicitly aim to generate sample reflecting general population |
| 10.1001/jama.2020.14765 | USA | Targets high-risk participants / Does not explicitly aim to generate sample reflecting general population |
| 10.1016/j.isci.2021.102489 | USA | US study without adjustment for race or ethnicity |
| https://jamanetwork.com/journals/jamanetworkopen/fullarticle/2777502?utm_source=For_The_Media&utm_medium=referral&utm_campaign=ftm_links&utm_term=031621 | USA | US study without adjustment for race or ethnicity |
| 10.1101/2021.01.27.21250615 | USA | US study without adjustment for race or ethnicity |
| 10.1101/2020.09.09.20191296v1 | USA | US study without adjustment for race or ethnicity |
| 10.3390/vaccines9050504 | Greece | Seroprevalence confidence interval reaches zero |
| 10.1101/2021.08.10.21261777v1 | Denmark | Crude seroprevalence less than 1 - test specificity |
| https://www.sciencedirect.com/science/article/pii/S266677622100096X | Andorra | Duplicate |
| 10.1101/2020.06.08.20125179v5 | Belgium | Duplicate |
| 10.1038/s41598-021-92775-y | Brazil | Duplicate with excluded study (EPICOVID19) |
| https://bit.ly/Epicovid19BRfases1-3 | Brazil | Duplicate/overlapping (10.1016/S2214-109X(20)30387-9) |
| 10.1101/2020.08.10.20171942v1 | Brazil | Duplicate/overlapping (10.1016/S2214-109X(20)30387-9) |
| https://www.covid19immunitytaskforce.ca/wp- | Canada | Duplicate |

| | | |
|---|---|---|
| content/uploads/2021/01/COVID-19-October-Report-for-CITF.pdf | | |
| 10.1111/trf.16296 | Canada | Duplicate |
| 10.1007/s10654-020-00716-2 | France | Duplicate |
| 10.1093/ije/dyab110 | France | Duplicate |
| 10.1016/j.idnow.2020.12.007 | France | Duplicate |
| 10.1101/2021.11.14.21265758 | India | Duplicate |
| 10.1016/S2214-109X(20)30544-1 | India | Duplicate |
| https://www.istat.it/it/files//2020/08/ReportPrimiRisultatiIndagineSiero.pdf | Italy | Duplicate |
| 10.1016/j.isci.2021.102646 | Qatar | Duplicate |
| 10.1101/2021.03.11.21253142v1 | Spain | Duplicate |
| 10.1016/j.lanepe.2021.100098 | UK | Duplicate |
| https://www.ukbiobank.ac.uk/media/x0nd5sul/ukb_serologystudy_report_revised_6months_jan21.pdf | UK | Duplicate |
| 10.1101/2020.10.26.20219725v1.full.pdf | UK | Duplicate |
| 10.1126/scitranslmed.abh3826 | USA | Duplicate |
| 10.1101/2020.11.10.20215145 | USA | Duplicate |

Note. The duplicate records shown in this table are not a comprehensive list of duplicates found.

**Appendix Table 3b.** Reports not included, sensitivity analysis: Including explicitly national-level general population studies without high risk of bias and with at least 500 participants aged ≥70 years.

| DOI or URL | Country/location | eligibility_revision_500_R1 |
|---|---|---|
| 10.1016/j.ejim.2021.01.029 | Vatican City | Conducted in 2021, identified at screening step |
| 10.1016/j.eclinm.2021.101172 | Zimbabwe | Conducted in 2021, identified at screening step |
| https://www.nicd.ac.za/wp-content/uploads/2021/03/COVID-19-Special-Public-Health-Surveillance-Bulletin-9-12-March-2021_.pdf | South Africa | Conducted in 2021, identified at screening step |
| 10.3201/eid2712.211465 | South Africa | Conducted in 2021, identified at screening step |
| 10.1101/2021.06.22.21258711 | Poland | Conducted in 2021, identified at screening step |
| 10.1101/2021.10.19.21265219v1.full-text | Peru | Conducted in 2021, identified at screening step |
| 10.1101/2021.03.23.21254169 | Iraq | Conducted in 2021, identified at screening step |
| 10.1101/2021.07.16.21260611v1.full-text | Bangladesh | Conducted in 2021, identified at screening step |
| 10.1101/2021.08.14.21262042 | Austria | Conducted in 2021, identified at screening step |
| 10.1093/cid/ciab626 | United States of America | Seroprevalence confidence interval reaches zero |
| 10.1101/2021.08.10.21261777v1 | Denmark | Crude seroprevalence less than 1 - test specificity |
| 10.1038/s41591-020-0949-6 | China | Duplicate, identified at screening step |
| 10.20452/pamw.15796 | Poland | Duplicate, identified at screening step |

| | | |
|---|---|---|
| https://www.theadvocate.com/baton_rouge/news/coronavirus/article_b499efa0-8983-11ea-a3f1-af936ecb1d2c.html | United States of America | Duplicate, identified at screening step |
| 10.1101/2020.05.01.20087478v1 | United States of America | Duplicate, identified at screening step |
| https://www.wbtv.com/2020/05/29/antibody-testing-continues-burke-county/&ct=ga&cd=CAAYGTIaOTBkZWE5ODk5NGU5MTg1OTpjb206ZW46VVM&usg=AFQjCNEvSO7bk5TLI9MT5-BeF8ILKdseXA | United States of America | Duplicate, identified at screening step |
| https://www.wcax.com/2020/07/17/early-results-of-uvm-research-tracking-covids-spread-in-vermont/&ct=ga&cd=CAAYCzIaOTBkZWE5ODk5NGU5MTg1OTpjb206ZW46VVM&usg=AFQjCNEblNmlKeakOFmkRkZMbbLnjhptxw | United States of America | Duplicate, identified at screening step |
| 10.3390/pathogens10060710 | United States of America | Duplicate, identified at screening step |
| 10.1177/00333549211055137 | United States of America | Duplicate, identified at screening step |
| https://wiadlek.pl/wp-content/uploads/archive/2021/WLek202105116.pdf | Ukraine | Duplicate, identified at screening step |
| 10.1128/JVI.01828-20 | Switzerland | Duplicate, identified at screening step |
| https://www.ne.ch/autorites/DFS/SCSP/medecin-cantonal/maladies-vaccinations/coronaimmunitas/Pages/R%c3%a9sultats-de-l'%c3%a9tude.aspx | Switzerland | Duplicate, identified at screening step |
| 10.1101/2021.07.31.21261428v2 | Russia | Duplicate, identified at screening step |
| 10.1016/S2214-109X(21)00386-7 | Ethiopia | Duplicate, identified at screening step |
| https://www.thelancet.com/journals/lanmic/article/PIIS2666-5247(20)30053-7/fulltext | China | Duplicate, identified at screening step |
| https://pitangueiras.sp.gov.br/wp-content/uploads/2020/12/Relatorio-Inquerito-COVID-19-Final.pdf | Brazil | Duplicate, identified at screening step |
| 10.1007/s12020-021-02728-8 | Brazil | Duplicate, identified at screening step |
| 10.2196/30406 | Brazil | Duplicate, identified at screening step |
| 10.1111/1753-6405.13155 | Australia | High risk of bias |
| 10.3201/eid2702.204088 | Japan | High risk of bias |
| https://www.haaretz.com/israel-news/.premium-up-to-270-000-israelis-had-coronavirus-new-study-concludes-1.8888435&ct=ga&cd=CAAYETIaOTBkZWE5ODk5NGU5MTg1OTpjb206ZW46VVM&usg=AFQjCNExqw1wlJNVBFrKKUd7hGTplMnR5A | Israel | High risk of bias |
| https://wellcomeopenresearch.org/articles/6-173 | Ghana | High risk of bias |
| 10.3390/pathogens10060774 | Croatia | High risk of bias |
| 10.1101/2020.09.09.20191296v1 | United States of America | High risk of bias |
| 10.1093/cid/ciab519 | United States of America | High risk of bias |
| 10.3390/v13081648 | Russia | COVID-19 mortality data unavailable for elderly at study location |
| 10.1101/2020.08.10.20171942v1 | Brazil | COVID-19 mortality data unavailable for elderly at study location |

| | | |
|---|---|---|
| 10.1016/j.ijid.2021.08.028 | Ethiopia | COVID-19 mortality data unavailable for elderly at study location |
| 10.1016/j.cmi.2021.06.002 | Iran | COVID-19 mortality data unavailable for elderly at study location |
| https://www.journals.vu.lt/AML/article/view/22344 | Lithuania | COVID-19 mortality data unavailable for elderly at study location |
| https://www.croiconference.org/abstract/us-population-based-survey-of-vaccine-willingness-and-sars-cov-2-antibody-prevalence/ | United States of America | No seroprevalence estimate for elderly |

Note. Excluded reports not listed that had not sampled ≥500 participants ≥70 years old; general population; or national-level.

**Appendix Table 4**. Seroprevalence estimate corrections for test performance using the Gladen-Rogan formula.

| First author last name | Assay sensitivity | Assay specificity | Seroprevalence description | Seroprevalence (%) | CI lower bound (%) | CI upper bound (%) | Gladen-Rogan corrected seroprevalence (%) | Gladen-Rogan corrected CI lower bound (%) | Gladen-Rogan corrected CI upper bound (%) |
|---|---|---|---|---|---|---|---|---|---|
| Abu-Raddad | 97.92 | 99.95 | 70-79 | 13.9 | | | 14.2 | | |
| Abu-Raddad | 97.92 | 99.95 | 80+ | 9.8 | | | 10.01 | | |
| Abu-Raddad | 97.92 | 99.95 | Adjusted, entire elderly group | | 11.5 | 14.7 | | 11.75 | 15.02 |
| Abu-Raddad | 97.92 | 99.95 | Crude, entire elderly group | 8.96 | 7.68 | 10.37 | 9.15 | 7.85 | 10.6 |
| Abu-Raddad | 97.92 | 99.95 | 0-9 | 5.5 | | | 5.62 | | |
| Abu-Raddad | 97.92 | 99.95 | 10-19 | 6.7 | | | 6.85 | | |
| Abu-Raddad | 97.92 | 99.95 | 20-29 | 23.1 | | | 23.6 | | |
| Abu-Raddad | 97.92 | 99.95 | 30-39 | 25.3 | | | 25.85 | | |
| Abu-Raddad | 97.92 | 99.95 | 40-49 | 30.1 | | | 30.75 | | |
| Abu-Raddad | 97.92 | 99.95 | 50-59 | 30.4 | | | 31.06 | | |
| Abu-Raddad | 97.92 | 99.95 | 60-69 | 23.9 | | | 24.42 | | |
| Anand | 100 | 99.8 | 65-79 | 8.3 | 7.8 | 8.9 | 8.31 | 7.82 | 8.92 |
| Anand | 100 | 99.8 | 80+ | 7.4 | 6.5 | 8.5 | 7.41 | 6.51 | 8.52 |
| Anand | 100 | 99.8 | Crude, entire elderly group | 7.64 | | | 7.65 | | |
| Anand | 100 | 99.8 | 18-44 | 9.8 | | | 9.82 | | |
| Anand | 100 | 99.8 | 45-64 | 9.5 | | | 9.52 | | |
| Charlton | 66.7 | 100 | 70-79 | 1.19 | 0.69 | 1.7 | 1.79 | 1.03 | 2.55 |
| Charlton | 66.7 | 100 | 80+ | 0.76 | 0.23 | 1.28 | 1.13 | 0.35 | 1.91 |
| Charlton | 66.7 | 100 | Crude, entire elderly group | 1.03 | | | 1.54 | | |
| Charlton | 66.7 | 100 | 0-9 | 2.52 | | | 3.78 | | |
| Charlton | 66.7 | 100 | 10-19 | 1.87 | | | 2.81 | | |
| Charlton | 66.7 | 100 | 20-29 | 3.38 | | | 5.07 | | |
| Charlton | 66.7 | 100 | 30-39 | 3.32 | | | 4.98 | | |
| Charlton | 66.7 | 100 | 40-49 | 2.16 | | | 3.25 | | |
| Charlton | 66.7 | 100 | 50-59 | 2.14 | | | 3.21 | | |
| Charlton | 66.7 | 100 | 60-69 | 1.47 | | | 2.2 | | |
| ISCII | 89.7 | 100 | 70-74 | 7.6 | 6.4 | 9.1 | 8.47 | 7.13 | 10.14 |
| ISCII | 89.7 | 100 | 75-79 | 7 | 5.6 | 8.7 | 7.8 | 6.24 | 9.7 |
| ISCII | 89.7 | 100 | 80-84 | 6.9 | 5.2 | 9.2 | 7.69 | 5.8 | 10.26 |
| ISCII | 89.7 | 100 | 85-89 | 6.6 | 4.5 | 9.5 | 7.36 | 5.02 | 10.59 |
| ISCII | 89.7 | 100 | 90+ | 6.5 | 3.9 | 10.8 | 7.25 | 4.35 | 12.04 |
| ISCII | 89.7 | 100 | 0-4 | 4.3 | | | 4.79 | | |
| ISCII | 89.7 | 100 | 5-9 | 5.9 | | | 6.58 | | |
| ISCII | 89.7 | 100 | 10-14 | 6.8 | | | 7.58 | | |
| ISCII | 89.7 | 100 | 15-19 | 6.1 | | | 6.8 | | |

| | | | | | | | | | |
|---|---|---|---|---|---|---|---|---|---|
| ISCII | 89.7 | 100 | 20-24 | 8.1 | | | 9.03 | | |
| ISCII | 89.7 | 100 | 25-29 | 7.6 | | | 8.47 | | |
| ISCII | 89.7 | 100 | 30-34 | 7 | | | 7.8 | | |
| ISCII | 89.7 | 100 | 35-39 | 6.7 | | | 7.47 | | |
| ISCII | 89.7 | 100 | 40-44 | 6.6 | | | 7.36 | | |
| ISCII | 89.7 | 100 | 45-49 | 7.3 | | | 8.14 | | |
| ISCII | 89.7 | 100 | 50-54 | 8.5 | | | 9.48 | | |
| ISCII | 89.7 | 100 | 55-59 | 7.3 | | | 8.14 | | |
| ISCII | 89.7 | 100 | 60-64 | 7.8 | | | 8.7 | | |
| ISCII | 89.7 | 100 | 65-69 | 7 | | | 7.8 | | |
| Merkely | 66.7 | 100 | Adjusted, entire elderly group | 0.75 | 0.21 | 1.29 | 1.12 | 0.31 | 1.93 |
| Merkely | 66.7 | 100 | Crude, entire elderly group | 0.62 | | | 0.93 | | |
| Merkely | 66.7 | 100 | 14-39 | 0.56 | | | 0.84 | | |
| Merkely | 66.7 | 100 | 40-64 | 0.7 | | | 1.05 | | |
| Paulino-Ramirez | 56.7 | 100 | Crude, entire elderly group | 5.97 | 5.13 | 6.94 | 10.53 | 9.05 | 12.24 |
| Paulino-Ramirez | 56.7 | 100 | 0-9 | 8.64 | | | 15.24 | | |
| Paulino-Ramirez | 56.7 | 100 | 10-19 | 3.85 | | | 6.79 | | |
| Paulino-Ramirez | 56.7 | 100 | 20-29 | 4.6 | | | 8.11 | | |
| Paulino-Ramirez | 56.7 | 100 | 30-39 | 5.09 | | | 8.98 | | |
| Paulino-Ramirez | 56.7 | 100 | 40-49 | 5.92 | | | 10.44 | | |
| Paulino-Ramirez | 56.7 | 100 | 50-59 | 6.09 | | | 10.74 | | |
| Royo-Cebrecos | 90.6 | 99.2 | Crude, entire elderly group | 13.41 | 12.41 | 14.46 | 14.92 | 13.82 | 16.1 |
| Warszawski, INSERM | 80 | 99.6 | 65-74 | 4.3 | 3.8 | 4.9 | 5.4 | 4.77 | 6.16 |
| Warszawski, INSERM | 80 | 99.6 | 75+ | 3.7 | 2.9 | 4.7 | 4.64 | 3.64 | 5.9 |
| Warszawski, INSERM | 80 | 99.6 | Crude, entire elderly group | 4.2 | | | 5.27 | | |
| Warszawski, INSERM | 80 | 99.6 | 15-17 | 9.8 | | | 12.31 | | |
| Warszawski, INSERM | 80 | 99.6 | 25-34 | 7.2 | | | 9.04 | | |
| Warszawski, INSERM | 80 | 99.6 | 35-44 | 6.5 | | | 8.16 | | |
| Warszawski, INSERM | 80 | 99.6 | 45-54 | 6.5 | | | 8.16 | | |
| Warszawski, INSERM | 80 | 99.6 | 55-64 | 5.3 | | | 6.65 | | |

**Appendix Table 5.** Uncorrected and seroreversion-corrected infection fatality rate in community-dwelling elderly

| Location (First author) | Antibody types measured | Study midpoint * | Peak of wave 1† | Time lag between primary date and peak (months) | IFR in community-dwelling elderly (uncorrected) | IFR in community-dwelling elderly (corrected for unmeasured antibody types) | IFR in community-dwelling elderly, corrected for 1% relative seroreversion/month‡ | IFR in community-dwelling elderly, corrected for 5% relative seroreversion/month‡ | IFR in community-dwelling elderly, corrected for 10% relative seroreversion/month‡ |
|---|---|---|---|---|---|---|---|---|---|
| Andorra (Royo-Cebrecos)** | IgG/IgM | May 23 | 2020-04-01 | 1.71 | 2.02 | 1.84 | 1.81 | 1.68 | 1.53 |
| Belgium (Herzog) | IgG | April 9 | 2020-04-07 | 0.07 | 3.79 | 3.13 | 3.13 | 3.12 | 3.11 |
| Canada (Saeed, Canadian Blood Services) | IgG | June 20 | 2020-04-30 | 1.68 | 3.23 | 2.67 | 2.62 | 2.45 | 2.24 |
| Canada (Ab-C Study Investigators) | IgG | July 23 | 2020-04-30 | 2.76 | 1.14 | 0.94 | 0.92 | 0.82 | 0.7 |
| Alberta, Canada (Charlton)** | IgG | December 15 | 2020-04-13 (2020-12-27) †† | 8.08 (-0.4) †† | 4.8 | 3.97 | 3.66 (3.97) †† | 2.62 (3.97) †† | 1.69 (3.97) †† |
| Ontario, Canada (Public Health Ontario, COVID-19 Immunity Task Force) | IgG | June 24 | 2020-04-26 | 1.94 | 1.96 | 1.62 | 1.59 | 1.47 | 1.32 |
| Denmark (Espenhain) | IgG/IgM/IgA | September 26 | 2020-03-31 | 5.88 | 2.92 | 2.92 | 2.75 | 2.16 | 1.57 |
| Denmark (Pedersen) | IgG/IgM/IgA | June 17 | 2020-03-31 | 2.56 | 4.43 | 4.43 | 4.32 | 3.89 | 3.38 |
| Dominican Republic (Paulino-Ramirez)** | IgG | May 22 | 2020-04-05 | 1.54 | 0.19 | 0.16 | 0.16 | 0.15 | 0.14 |
| France (Warszawski, INSERM)** | IgG | December 1 | 2020-11-11 | 0.66 | 4.14 | 3.42 | 3.4 | 3.31 | 3.19 |
| Ile-de-France, France (Carrat) | IgG | May 21 | 2020-03-29 | 1.74 | 7.1 | 5.87 | 5.77 | 5.37 | 4.89 |
| Nouvelle-Aquitaine, France (Carrat) | IgG | May 21 | 2020-03-29 | 1.74 | 1.27 | 1.05 | 1.03 | 0.96 | 0.87 |
| Hungary (Merkely)** | IgG | May 15 | 2020-04-16 | 0.95 | 1.85 | 1.53 | 1.51 | 1.45 | 1.38 |

| | | | | | | | | | |
|---|---|---|---|---|---|---|---|---|---|
| Iceland (Gudbjartsson) | IgG/IgM/IgA | June 1 | 2020-04-02 | 1.97 | 3.12 | 3.12 | 3.06 | 2.82 | 2.53 |
| India (Murhekar) | IgG | September 11 | 2020-09-10 | 0.03 | 0.43 | 0.36 | 0.36 | 0.36 | 0.36 |
| Tamil Nadu, India (Malani) | IgG | November 16 | 2020-07-18 | 3.98 | 0.32 | 0.27 | 0.26 | 0.22 | 0.18 |
| Italy (ISTAT) | IgG | June 26 | 2020-03-26 | 3.02 | 7.63 | 6.31 | 6.12 | 5.4 | 4.59 |
| Qatar (Abu-Raddad)** | IgG | July 5 | 2020-06-23 | 0.39 | 2.18 | 1.8 | 1.79 | 1.76 | 1.73 |
| Spain (ISCII)** | IgG | November 29 | 2020-03-27 | 8.11 | 4.55 | 3.76 | 3.46 | 2.48 | 1.6 |
| UK (NA) | Missing/Unclear | July 12 | 2020-04-06 | 3.19 | 3.53 | 3.53 | 3.42 | 3 | 2.52 |
| England (Ward) | IgG | July 9 | 2020-04-03 | 3.19 | 9.68 | 8 | 7.75 | 6.8 | 5.72 |
| England and Wales (Public Health England) | Missing/Unclear | May 22 | 2020-04-03 | 1.61 | 7.55 | 7.55 | 7.43 | 6.95 | 6.37 |
| Greater Glasgow and Clyde, Scotland (Hughes) | IgG | April 26 | 2020-04-13 | 0.43 | 2.87 | 2.37 | 2.36 | 2.32 | 2.27 |
| USA (Anand)** | IgG/IgM/IgA | July 14 | 2020-04-14 | 2.99 | 1.2 | 1.2 | 1.17 | 1.03 | 0.88 |
| USA (Kalish) | IgG/IgM/IgA | June 27 | 2020-04-14 | 2.43 | 2.27 | 2.27 | 2.22 | 2 | 1.76 |
| Netherlands (Vos) | IgG | June 21 | 2020-04-01 | 2.66 | 2.27 | 1.88 | 1.83 | 1.64 | 1.42 |

* Midpoint of the seroprevalence sampling period. † The peak of the first epidemic wave was defined as one week before the date with the highest rolling average 7-day mortality (according to Worldometer, situational reports, or Wikipedia). The first epidemic wave was defined to end by the date with the lowest 7-day average of daily deaths since the beginning of the epidemic. If two or more dates were tied for peak values, we chose the date corresponding to the midpoint between the first and last one. ‡ Seroreversion correction of the IFR by $X^m$-fold, where m is the number of months from the peak of the first epidemic wave in the specific location and X is given values of 0.99, 0.95, and 0.90 corresponding to 1%, 5%, and 10% relative rate of seroreversion every month, respectively.** Seroprevalence corrected for test performance with the Gladen-Rogan formula. †† For Alberta, Canada, main estimates are based on the peak of the first wave; estimates based on peak of the second wave are given within parentheses.

**Appendix Table 6**. Sensitivity analysis with a later cutoff for cumulative COVID-19 mortality (study midpoint plus two weeks instead of plus one week).

| **Location (First author)** | **Closest date to study midpoint + 1 week, with total deaths available** | **Closest date to study midpoint + 2 weeks, with total deaths available** | ***Relative* increase in IFR from cutoff +1 week to cutoff +2 weeks after study midpoint (%)** | **Corrected IFR in community-dwelling elderly, using study midpoint + 1 week (%)** | **Corrected IFR in community-dwelling elderly, using study midpoint + 2 weeks (%)** |
|---|---|---|---|---|---|
| Andorra (Royo-Cebrecos)** | 2020-05-23 | 2020-05-30 | 0 | 1.84 | 1.84 |
| Belgium (Herzog) | 2020-04-09 | 2020-04-16 | 56.51 | 3.13 | 4.9 |
| Canada (Saeed, Canadian Blood Services) | 2020-06-14 | 2020-06-29 | 5.16 | 2.67 | 2.81 |
| Canada (Ab-C Study Investigators) | 2020-07-28 | 2020-07-28 | 0 | 0.94 | 0.94 |
| Alberta, Canada (Charlton)** | 2020-12-15 | 2020-12-22 | 5.85 | 3.97 | 4.2 |
| Ontario, Canada (Public Health Ontario, COVID-19 Immunity Task Force) | 2020-06-24 | 2020-07-01 | 1.71 | 1.62 | 1.65 |
| Denmark (Espenhain) | 2020-09-25 | 2020-10-02 | 1.4 | 2.92 | 2.96 |
| Denmark (Pedersen) | 2020-06-22 | 2020-06-24 | 0.84 | 4.43 | 4.47 |
| Dominican Republic (Paulino-Ramirez)** | 2020-05-22 | 2020-05-29 | 7.02 | 0.16 | 0.17 |
| France (Warszawski, INSERM)** | 2020-12-02 | 2020-12-09 | 5.32 | 3.42 | 3.6 |
| Ile-de-France, France (Carrat) | 2020-05-26 | 2020-05-28 | | 5.87 | 5.87 |
| Nouvelle-Aquitaine, France (Carrat) | 2020-05-25 | 2020-05-28 | | 1.05 | 1.05 |
| Hungary (Merkely)** | 2020-05-15 | 2020-05-22 | 7.69 | 1.53 | 1.64 |
| Iceland (Gudbjartsson) | 2020-06-01 | 2020-06-08 | 0 | 3.12 | 3.12 |
| India (Murhekar) | 2020-11-20 | 2020-09-18 | 10.46 | 0.36 | 0.4 |
| Tamil Nadu, India (Malani) | 2020-11-20 | 2020-11-23 | | 0.27 | 0.27 |
| Italy (ISTAT) | 2020-06-26 | 2020-07-03 | 0.34 | 6.31 | 6.33 |
| Qatar (Abu-Raddad)** | 2020-07-05 | 2020-07-12 | 14.84 | 1.8 | 2.07 |
| Spain (ISCII)** | 2020-11-29 | 2020-12-06 | 3.13 | 3.76 | 3.88 |

| | | | | | |
|---|---|---|---|---|---|
| UK (NA) | 2020-07-10 | 2020-07-19 | 0.37 | 3.53 | 3.55 |
| England (Ward) | 2020-07-10 | 2020-07-17 | 5.57 | 8 | 8.45 |
| England and Wales (Public Health England) | 2020-05-22 | 2020-05-29 | 3.92 | 7.55 | 7.85 |
| Greater Glasgow and Clyde, Scotland (Hughes) | 2020-04-26 | 2020-05-03 | 23.03 | 2.37 | 2.92 |
| USA (Anand)** | 2020-07-14 | 2020-07-21 | 4.27 | 1.2 | 1.26 |
| USA (Kalish) | 2020-06-27 | 2020-07-04 | 2.84 | 2.27 | 2.34 |

** Seroprevalence corrected for test performance with the Gladen-Rogan formula.

### Appendix text

Most of those studies reporting IFR in the elderly represent locations with the highest reported IFRs for the overall population, thus the reported IFR estimates for the elderly may also be substantially inflated versus the global experience. For example, Pastor-Barriuso *et al* (1) used nationwide estimates from Spain (2) (ENE-COVID study, comprised by our sample (3)) to calculate the IFR in community-dwelling persons, reporting an IFR in individuals 70-79 years old of 4.96% and in those 80 years or older of 11.6% using confirmed COVID-19 deaths. The estimates did not exclude deaths of nursing home residents that occurred in hospitals. Molenberghs *et al* (4) used a statistical model based on seroprevalence (5) and granular national data from Belgium to estimate IFRs for the community-dwelling at 1.2% for individuals 70-79 years, 1.0% for 80-89 years, and 2.4% for 90 years or older (slightly lower than our estimates). Blackburn *et al* (6) calculated the IFR in Indiana, USA, at 1.71% for community-dwelling persons aged 60 years or older. The seroprevalence study used as basis for these calculations (7) was later updated by the same team using statistical methods to address "nonresponse among various demographic groups and to adjust for testing errors to reduce bias in the estimates of the overall disease prevalence", resulting in an upweighting of individuals identifying as Black or "Other (including multiracial)" non-white, or with Hispanic ethnicity, and an overall seroprevalence of 3.6% instead of 2.8% (8). Overall, the Indiana results are quite consistent with our estimates. Conversely, Mahajan *et al* (9) report the IFR in Connecticut, USA, at 16.46% for community-dwelling persons 65 years or older. This estimate depends on seroprevalence data for very few people >65 (n=187).
